# NINM Safety Parameter Explorer: a safety-reference tool for non-invasive neuromodulation

**DOI:** 10.64898/2026.09.27.26364117

**Authors:** Yossi Zana, Isabela Rocha Fernandes, Tiago da Silva Lopes

**Author notes:** **Corresponding author:** Yossi Zana.

## Abstract

**Introduction:** Safety parameters for non-invasive neuromodulation (NINM) remain scattered across consensus documents that differ in format, units, and scope, so pre-session parameter screening stays error-prone and difficult to audit.

**Objectives:** To describe the NINM Safety Parameter Explorer, a browser-based tool that consolidates safety thresholds for six NINM modalities (e.g., TMS, tDCS, peripheral magnetic stimulation, TENS, transcranial ultrasound, and transcutaneous vagus nerve stimulation) into a single rule engine in which every numeric boundary carries an explicit, auditable provenance classification as literature-derived, literature-anchored, or operational default.

**Methods and Materials:** We encoded the tool’s rule corpus from published international consensus guidelines and peer-reviewed literature (82 modality threshold rules: 29 literature-derived, 36 literature-anchored, 17 operational defaults; complemented by 5 target-site caps for 87 total evaluative rules, alongside 15 informational reference values), then evaluated it in a simulated, automated internal verification suite of 38 example protocols spanning all six modalities and three expected verdicts, with automated static integrity assertions, regression guards, and behavioural and accessibility unit checks, all under a single pinned software version (v1.1.0-beta).

**Results:** All 38 examples agreed with their declared verdicts (19 safe, 8 caution, 11 unsafe), and all static guards and supporting checks passed.

**Conclusion:** The Explorer shows complete internal self-consistency between its rule corpus, engine, and interface, and its per-rule provenance labelling renders safety reasoning transparent and auditable for research and educational use. The tool is an in-silico protocol planning aid; it is not a medical device and is not intended for clinical decision-making or use in humans.

## Introduction

Non-invasive neuromodulation (NINM) has matured into a routine clinical and laboratory modality, encompassing repetitive transcranial magnetic stimulation (rTMS) and accelerated theta burst stimulation (TBS) [1–5]. The field further includes transcranial electrical stimulation (tES; comprising tDCS, tACS, and tRNS) [6, 7], transcranial ultrasound stimulation (tUS) [8–11], and peripheral approaches such as transcutaneous vagus nerve stimulation (tVNS) and peripheral magnetic stimulation [12–14]. Across these diverse modalities, clinical teams must navigate a fragmented landscape of multi-document consensus guidelines to complete pre-session safety screening.

Each modality carries distinct, non-interchangeable quantitative safety conventions. For rTMS, seizure induction risk is governed by stimulation frequency, train duration, cumulative pulse counts, and concomitant pharmacotherapy, while acoustic output mandates hearing protection [15–17]. For tES, operative limits are cutaneous and electrochemical [18, 19]; repeated daily tDCS at electrode current densities of about 0.06 mA/cm² (25–35 cm² electrodes; 1.5–2.1 mA) has caused persisting skin lesions, while the rat-cortical charge-density threshold of 5.24 C/cm² marks a boundary no protocol should target [20], with feasibility supported by escalation studies [21]. For tUS, exposure limits draw on indices from diagnostic ultrasound (mechanical index 1.9, thermal index 6, I_SPTA_ 720 mW/cm^2^, and I_SPPA_ 190 W/cm^2^) [22, 23], refined by neuromodulatory consensuses [10, 11] and parameter reviews [24, 25]. Peripheral stimulation safety relies on device ceilings and clinical parameter ranges [13, 14, 26, 27]. For tVNS, heterogeneous parameter reporting and autonomic considerations constrain protocol design [12, 28–31]. Concomitant pharmacotherapy reshapes these envelopes entirely, since medications can lower seizure thresholds or alter responsiveness to rTMS and tES [17, 32, 33].

These conventions reach the laboratory as multi-parameter tables scattered across consensus documents that differ in format, units, and scope. Confirming that a planned protocol respects every relevant boundary therefore requires manual cross-referencing and unit conversion, which remains a persistent operational vulnerability. Documented failure modes include catastrophic unit mismatches, such as treating Bikson et al.’s [18] 7.2 C session limit as 7.2 C/cm², a multi-fold error. Manual lookups of tabular literature (such as Rossi et al. [16] Table 4) are prone to alignment errors. Pre-session screening is currently distributed across four classes of instruments, each of them incomplete in its own way. Screening questionnaires [34] and their parent consensus documents [15–17] filter subject-level contraindications but remain blind to specific planned parameter sets and medication interactions. Computational field-modeling suites (such as SimNIBS and ROAST) deliver individualized spatial dosimetry yet presuppose structural MRI and substantial offline computation, and they carry no encoded consensus boundaries. Stimulator firmware interlocks protect hardware rather than evaluating protocol combinations against consensus limits, and unvalidated spreadsheets remain prone to unit-conversion errors. This gap widens as clinical teams increasingly deploy multi-modal experimental protocols, forcing investigators to reconcile incompatible unit conventions and divergent screening forms across disconnected spreadsheets.

To close this gap, we developed the Non-invasive Neuromodulation Safety Parameter Explorer (NINM Safety Explorer), a free, zero-install, client-side web application (https://neuromod.pesquisa.ufabc.edu.br/NINM_Safety/) that evaluates planned protocols before clinical or laboratory commitment. It covers six modalities: transcranial magnetic stimulation (TMS)/TBS, transcranial electrical stimulation (specifically tDCS in the current software release), peripheral magnetic stimulation (PMS), transcutaneous electrical nerve stimulation (TENS), transcranial ultrasound stimulation (tUS), and tVNS. This breadth is achieved without sacrificing depth, since each modality operates through an isolated, specialized evaluator that executes full dosimetric derivations and consensus rules (e.g., cell-for-cell Rossi Table 4 for rTMS, electrode-area-scaled charge density for tDCS, and duration-scheduled thermal indices for tUS).

The system comprises five modules: (i) rule-based parameter evaluation encoding consensus boundaries for each modality; (ii) automated computation of derived quantities (e.g., current density, cumulative charge, duty cycles); (iii) a pharmacological risk engine screening for compounds altering stimulation response; (iv) interactive two-dimensional safety-envelope visualization placing a planned protocol relative to published boundaries; and (v) auditable protocol reporting recording provenance tiers. The knowledge base is structured across a provenance taxonomy: Tier 1 comprises literature-derived consensus limits, Tier 2 encodes literature-anchored observation baselines, Tier 3 applies operational defaults, and Category 4 provides non-evaluative informational reference values.

The Explorer is a research and educational instrument, not a medical device or clinical decision support, and provides no clearance to stimulate a human subject. Its contributions are fivefold: (1) a unified rule engine spanning six modalities; (2) automated unit arithmetic and derived-quantity computation; (3) structured medication–protocol interaction screening; (4) visual proximity feedback on published boundaries; and (5) exportable audit trails suitable for laboratory records. The remainder of the paper describes the system and rule encoding (Section 2, Methods), reports the automated internal verification suite (Section 3, Results), and discusses the scope, limitations, and intended use of the tool (Section 4).

## Methods

### System overview and design constraints

The Explorer is a client-side web application that evaluates a planned non-invasive neuromodulation protocol before it is committed to laboratory documentation. It covers six modalities, namely TMS, transcranial electrical stimulation, PMS, TENS, tUS, and tVNS, with transcranial electrical stimulation currently limited to tDCS in this software release, and for each submitted protocol it returns a verdict, per-parameter findings, contraindication and medication warnings, and an exportable report in which every verdict can be traced back to its source.

The entire application executes in the browser. Assessment logic, the threshold knowledge base, contraindication screening, visualization, and report generation all run locally, with no server-side computation and no network communication once the page has loaded. The deployed artifact is a static directory of 18 files that runs from any static host and, because nothing beyond the entry page is fetched, also directly from a laboratory machine’s file system with no installation step. This runtime model has consequences that were treated as design constraints rather than afterthoughts. Because subject-specific inputs (parameters, medical history, medication list) never leave the machine on which the tool runs, and no telemetry, analytics, or external resources are contacted, privacy here follows from the architecture itself rather than from policy. The deployment boundary is enforced structurally as well: the release assembly is generated from an explicit file manifest, so internal documentation and the copyrighted source library used during development are incapable of being published with the application. The tool’s research-tier status is likewise enforced in software. A session cannot begin until a non-dismissible acknowledgement is accepted, stating that the tool is a research and educational instrument, not a medical device, that it must not be used to clear a human stimulation procedure, and that responsibility rests with the investigator; the same statement is repeated beside every displayed result and inside every export.

### Model architecture

The system is organized in four layers (Fig. 1). A user-interface layer presents the input forms and results; an application controller collects and validates inputs, dispatches assessments, and renders outputs; a safety assessment engine holds the domain logic; and a reporting layer produces on-screen results and exports. Two properties of this architecture underpin everything else. The safety knowledge base acts as a single declaration point, meaning that every threshold in the system is declared in one component together with its citation registry and no other layer may assert a limit; this is what makes per-rule provenance mechanically auditable and lets the verification suite re-derive the safety envelope at every run. The assessment engine, in turn, is a pure function of its inputs, performing no input/output and holding no state between assessments, and it returns the same assessment for the same parameters. This determinism makes the internal verification of Section 3 meaningful and allows the engine to be tested independently of the interface.

**Figure 1:**
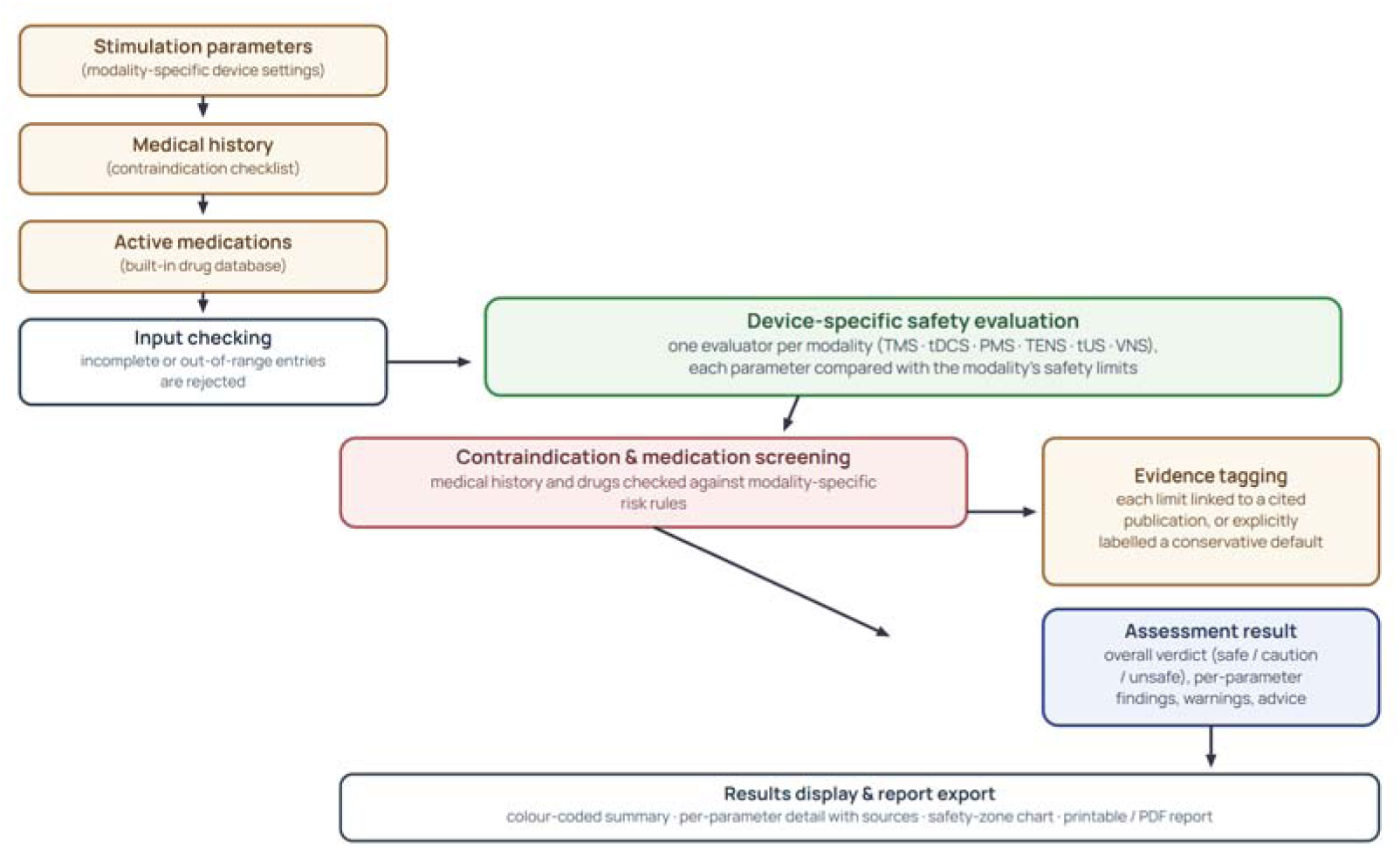
Assessment data flow. A submitted protocol passes through the input, controller, assessment-engine, and reporting layers; every reported finding is annotated with the threshold’s provenance tier: literature-derived, literature-anchored, or operational default.

### Inputs

Assessment rests on three input channels (Fig. 2): the planned stimulation parameter set, entered through modality-specific fields with native range checking, where incomplete or out-of-range entries are rejected before evaluation rather than silently coerced; a structured medical-history checklist covering contraindication-relevant conditions; and the active medication list, matched against a built-in drug database so that pharmacological risk screening requires no external lookup. The complete list of compounds and interaction classes encoded in this database is provided in Supplementary Table S2. Only these drugs are considered by the screening engine, and any medication outside this list is not evaluated and produces no pharmacological warning. For focused ultrasound (tUS), an essential physical boundary applies, since the Explorer evaluates in-situ acoustic exposure in target brain tissue in accordance with ITRUSST international consensus guidelines (Aubry et al. [10]). The application evaluates tissue exposure but does not perform skull transmission or numerical acoustic modeling; users must enter derated intracranial exposure parameters (, MI, TI) estimated from external simulation or empirical transmission models rather than free-field water-tank measurements. To support exploration and training, the tool ships a curated library of 38 fully parameterized example protocols spanning all six modalities (12 TMS, 5 tDCS, 5 PMS, 5 TENS, 5 tUS, 6 tVNS), banded at design time as safe (19), caution (8), or unsafe (11); selecting an example populates the form without producing a verdict, so a submitted assessment is always an explicit user action.

**Figure 2:**
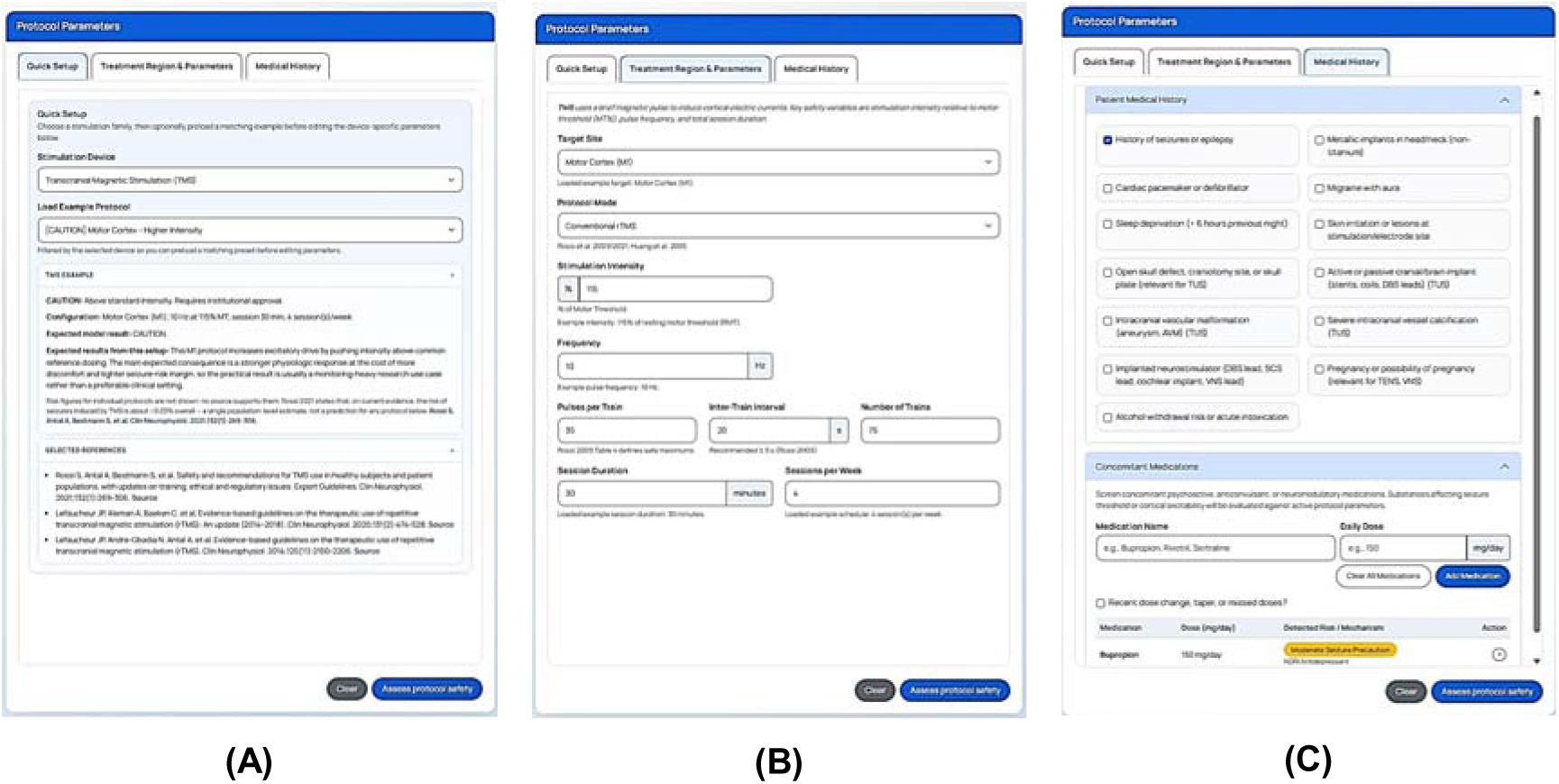
Protocol Parameters panel of the graphical user interface, with its three input tabs: (A) Quick Setup: stimulation-device selection and the curated example-protocol library; (B) Treatment Region & Parameters: modality-specific parameter entry; (C) Medical History: structured contraindication checklist and active medication list.

### Knowledge base curation, inclusion criteria, and extraction methodology

The safety thresholds and operational rules encoded in the Explorer were curated through a systematic, multi-stage evidence synthesis pipeline that extracts quantitative safety boundaries from indexed biomedical literature and international consensus statements.

#### Search Strategy and Bibliographic Sources

Bibliographic queries targeting the six modalities were executed across indexed databases (PubMed/MEDLINE, Scopus, IEEE Xplore, Google Scholar) and archives of international standardization consortia (IFCN, ITRUSST, IEC) through September 2026. Full search strings combining controlled vocabulary and safety-specific limiters are available in Supplementary Materials.

#### Hierarchical Evidence Framework

Extracted evidence was stratified across a three-level hierarchy to establish defensible safety limits. Level 1 prioritized official international consensus guidelines governing TMS [1, 2, 16, 17], tUS [10, 11], and tES/tDCS [6, 7, 18, 20]. Level 2 incorporated large-scale systematic reviews and meta-analyses establishing empirical parameter distributions for auricular tVNS [29], cervical tVNS [28], TENS [13, 27], PMS [14], and tUS [22, 24, 25]. Level 3 included canonical protocol descriptions defining standard pulse geometries [3] and landmark human tUS protocols [8], along with biophysical damage ceilings.

#### Inclusion and Exclusion Criteria

Inclusion required publication in a peer-reviewed journal or official standard, reporting of quantitative limits, and direct application to human non-invasive neuromodulation. Conflicting bounds were resolved by prioritizing the highest-ranking consensus standard. Exclusion criteria disqualified unvetted preprints, commercial materials, un-derated free-field tUS measurements, and animal studies. One necessary exception was made for the 5.24 C/cm² rat cortical lesion threshold reviewed in [20], serving as an essential biological ceiling since human tissue lesion induction is ethically precluded.

#### Rectification of Material Transcription and Attribution Errors

The dual-pass audit rectified several transcription errors common in legacy neuromodulation tools. A widely cited legacy boundary of 7.2 C/cm² was identified as a units conflation: 7.2 C represents a total session charge envelope [18], whereas 5.24 C/cm² marks the biological tissue lesion limit [20]. A re-extraction of Table 4 in [16] restored fidelity to the 25 distinct cells, explicitly tagging the 13 cells representing longest tested durations rather than demonstrated seizure ceilings [16]. The 30 mA TENS boundary in [13] was reclassified as an analgesic dose floor rather than an adverse-event ceiling. Throughout, secondary provenance was maintained by explicitly citing primary experimental findings as adopted by international consensus reviews [16, 20, 33].

### Conflict resolution

#### Deterministic Rule Encoding and Dosimetric Arithmetic

The deterministic rule engine integrates with a clinician’s pre-session workflow by computing derived dosimetric quantities, such as charge density and duty-cycle geometry, and comparing them against modality-specific safety boundaries. Malformed numeric input is strictly rejected rather than defaulted.

#### Resolution of Conflicting Literature

Where sources provided conflicting parameter ceilings, arbitration followed chronological supersession and biophysical fidelity. The canonical cases structuring the knowledge base are these:

1. *IFCN consensus safety grid vs. exploratory data [TMS] [15, 16]:* The Explorer adopted Table 4 from [16] as the superseding standard over exploratory historical data [15], constraining evaluations to the 90-130% RMT grid.
2. *IFCN consensus reaffirmation [TMS] [16, 17]:* Following the update in [17] which reaffirmed earlier guidelines, the engine directly encodes Table 4 of [16] as the persistent reference standard, in concordance with [17].
3. *ITRUSST vs. FDA Diagnostic Ultrasound [tUS] [10, 11, 23]:* Prioritizing ITRUSST consensus [10, 11] over FDA diagnostic ceilings [23], thermal risk is evaluated through ratified thermal dose metrics rather than _SPTA_, which was retained strictly for advisory context.
4. *Clinical trial envelopes vs. biological tissue lesion limits [tDCS] [18, 20]:* The system integrates clinical trial envelopes (7.2 C cumulative charge) [18] with specific tissue lesion limits (0.06 mA/cm² skin boundary and 5.24 C/cm² biological ceiling) [20], evaluating session charge against the envelope in [18].

#### Evidence Tier Classification Rubric

To prevent exploratory trial parameters from being misattributed as formal safety ceilings, every rule was assigned to a three-tier provenance taxonomy that renders technical specifications clinically readable:

1. Tier 1: Consensus trial limits (literature-derived). Upper safety boundaries and stopping rules from official international consensus guidelines, resolved through the application’s citation registry.
2. Tier 2: Evidence-extrapolated limits (literature-anchored). Parameters derived from published observational studies or clinical experience, serving as adverse-event observation points rather than absolute ceilings.
3. Tier 3: Precautionary boundaries (operational defaults). Engineering baselines and conservative caps established where peer-reviewed literature is silent or ambiguous.

Additionally, non-evaluative properties, including diagnostic benchmarks like the FDA diagnostic ultrasound _SPPA_ ceiling of 190 W/cm² [23], are segregated into an informational reference values class (Category 4), providing essential advisory context without participating in safety verdicts.

The same provenance objects render in the interface, the print view, and every export, so an assessment remains auditable outside the application.

### Rule inventory and traceability

The knowledge base was enumerated mechanically from its single declaration point through a context-free parameter extraction of the threshold module in which every parameter was classified by the engine’s own provenance function, yielding the inventory summarized in Table 1 and reproduced rule by rule in Supplementary Table S1. The engine declares 82 modality threshold rules: 29 literature-derived, each resolved through the 36-entry citation registry; 36 literature-anchored, carrying the mandatory published-value non-limit marker; and 17 operational defaults carrying the explicit unsourced operational-default marker. Across the software UI, the reference registry contains 36 entries across six modality groups; within the active threshold knowledge base, 22 unique reference keys anchor rule evaluations, 13 of which directly support the 29 literature-derived rules (distinguished from the manuscript’s 34-entry bibliography, which additionally encompasses medication screening sources, diagnostic criteria, and general trial literature). Three special-case tables complement these declarations: the rTMS maximum train-duration table adapted cell-for-cell from Rossi et al. [16] (25 cells: five frequency bands by five stimulation intensities, thirteen of which the source marks “>”, meaning the longest duration tested rather than a demonstrated ceiling, and which the tool reports as such), and the two fixed theta-burst protocol definitions of Huang et al. [3]. Five target-site session-duration caps complete the evaluative safety rules (4 literature-anchored, 1 operational default), giving 87 evaluative safety rules in total. Fifteen informational reference values, twenty-one target-site display labels, and two citation metadata strings appear in Supplementary Table S1 for completeness and advisory context, but are non-evaluative and not counted as safety rules.

**Table 1:**
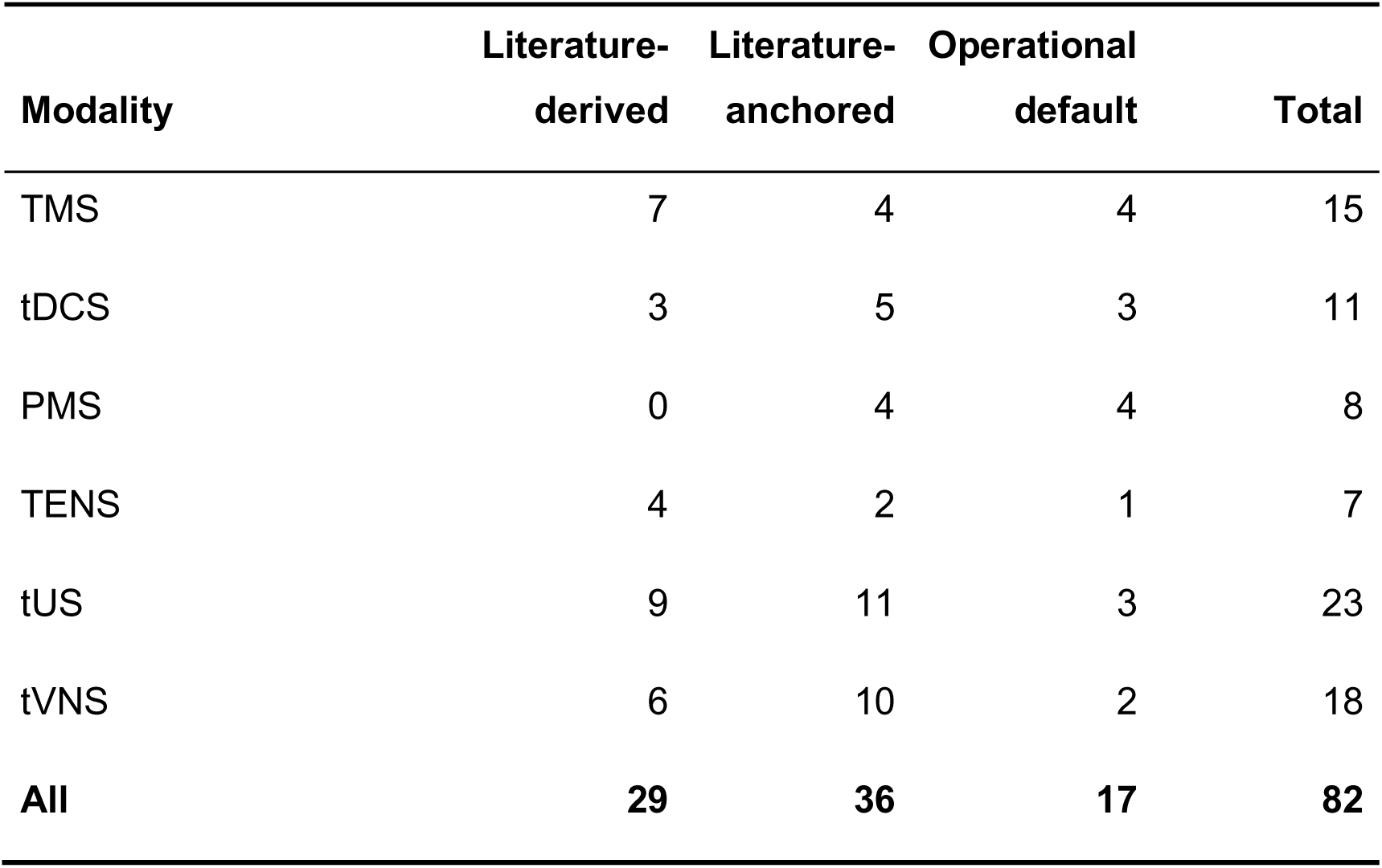
Rule inventory by modality. Counts are from a mechanical extraction of the knowledge base at the pinned version (v1.1.0-beta); the full rule-by-rule listing is Supplementary Table S1. Note: tDCS includes both common and caution current-density limits, which share the published 0.06 mA/cm² ceiling to enforce an immediate unsafe transition without an intermediate caution band.

The composition of this corpus is worth noting. Documented literature provenance (literature-derived plus literature-anchored) accounts for 65 of the 82 modality rules (79.3%), with operational defaults making up the remaining 17 rules (20.7%). Literature-derived rules anchor the principal limits across all modalities, whereas literature-anchored rules represent practice baselines such as the tDCS 0.06 mA/cm² current-density boundary [20], PMS train boundaries [14], TENS intensity ranges [13, 27], tUS acoustic frequencies [22, 25], and tVNS titration anchors [28, 29].

The 17 operational defaults comprise cumulative dosimetry ceilings and unstandardized intensity precautions (detailed fully in Supplementary Materials). Threshold boundaries and classifications were formally standardized in version v1.1.0-beta, resolving legacy parameter inversions. The remaining 15 non-evaluative properties are segregated as Category 4 informational reference values, including lower-bound practice envelopes and diagnostic references (e.g., FDA diagnostic ultrasound ceilings), providing advisory context without conflating the evaluative rule count.

### Contraindication and medication screening

To augment clinical safety checklists, a dedicated screening module evaluates twelve medical conditions (including metallic implants and seizure history) alongside concomitant pharmacotherapy for seizure-threshold interactions. These conditions are evaluated according to modality-specific biophysical interactions and international consensus guidelines, namely the IFCN guidelines of Rossi et al. [16, 17], the tES guidelines of Antal et al. [20], the ITRUSST guidelines of Murphy et al. [11] and Aubry et al. [10], and IEC 60601-2-10 [26]. The complete list is itemized in Supplementary Table S3.

Concomitant medication is evaluated against the 53-substance database (Supplementary Table S2). To prevent false positives from short brand names, user input is matched against canonical names and aliases using deterministic exact-string equality. The pharmacological knowledge base encodes proconvulsant compounds across the consensus hazard tiers of Rossi et al. [16]: List 1 strong hazards, List 2 relative hazards, and acute substance withdrawal vulnerabilities, listed as List 3 in Section 5.3 of the same guideline [16]. Unrecognized medications are assigned an explicit uncatalogued descriptor, requiring independent investigator review.

## Results interface and reporting

The assessment returns its feedback through an immediate traffic-light display, structured as an object containing an overall verdict, per-parameter findings, and contraindication warnings. Individual parameters are graded on a five-level evaluation scale:

1. Good: Nominal / unconstrained baseline parameter within conventional operating envelopes.
2. Safe: Parameter strictly conforms to established literature safety boundaries.
3. Caution: Parameter exceeds clinical practice baselines or enters an adverse-event observation zone governed by published clinical experience (Tier 2).
4. Warning: Parameter exceeds an operational engineering precaution or conservative default cap (Tier 3).
5. Unsafe: Parameter breaches an international consensus safety ceiling (Tier 1) or biological tissue damage limit.

The overall protocol assessment is computed deterministically, aggregating the individual parameter grades according to their tier:

- Absolute Contraindication Gate: If any medical-history condition or active compound evaluates to an absolute contraindication, the system immediately emits an Absolute Contraindication verdict, displaying a prominent critical banner that overrides numerical parameter clearance.
- Unsafe Verdict (Tier 1 Consensus Breach): If any individual parameter receives an Unsafe rating (violating a Tier 1 literature-derived consensus ceiling), the protocol is designated Unsafe (“Exceeds consensus safety limit: not permitted”).
- Caution Verdict (Tier 2 Practice Envelope): In the absence of an unsafe breach, if any parameter triggers a Caution or Warning status governed by Tier 2 literature-anchored trial evidence, the overall verdict evaluates to Caution (“Outside published clinical experience: specialist review”).
- Caution Verdict (Tier 3 Operational Precaution): If breaches are confined exclusively to Tier 3 operational defaults, the overall verdict evaluates to Caution (“Exceeds operational precaution: review accelerated pacing”).
- Safe Verdict: If all evaluated parameters evaluate to Safe or Good and no contraindications are present, the protocol evaluates to Safe (“Within conventional safety envelope”).

The results view (Fig. 3) comprises a colour-coded summary, expandable per-parameter details showing the applied threshold provenance, and an interactive safety-envelope chart. All user-facing text is bilingual (English/Portuguese), and session results are held purely in-memory; the sole durable outputs are explicitly generated exports, each permanently embedding the research-tier disclaimer.

**Figure 3:**
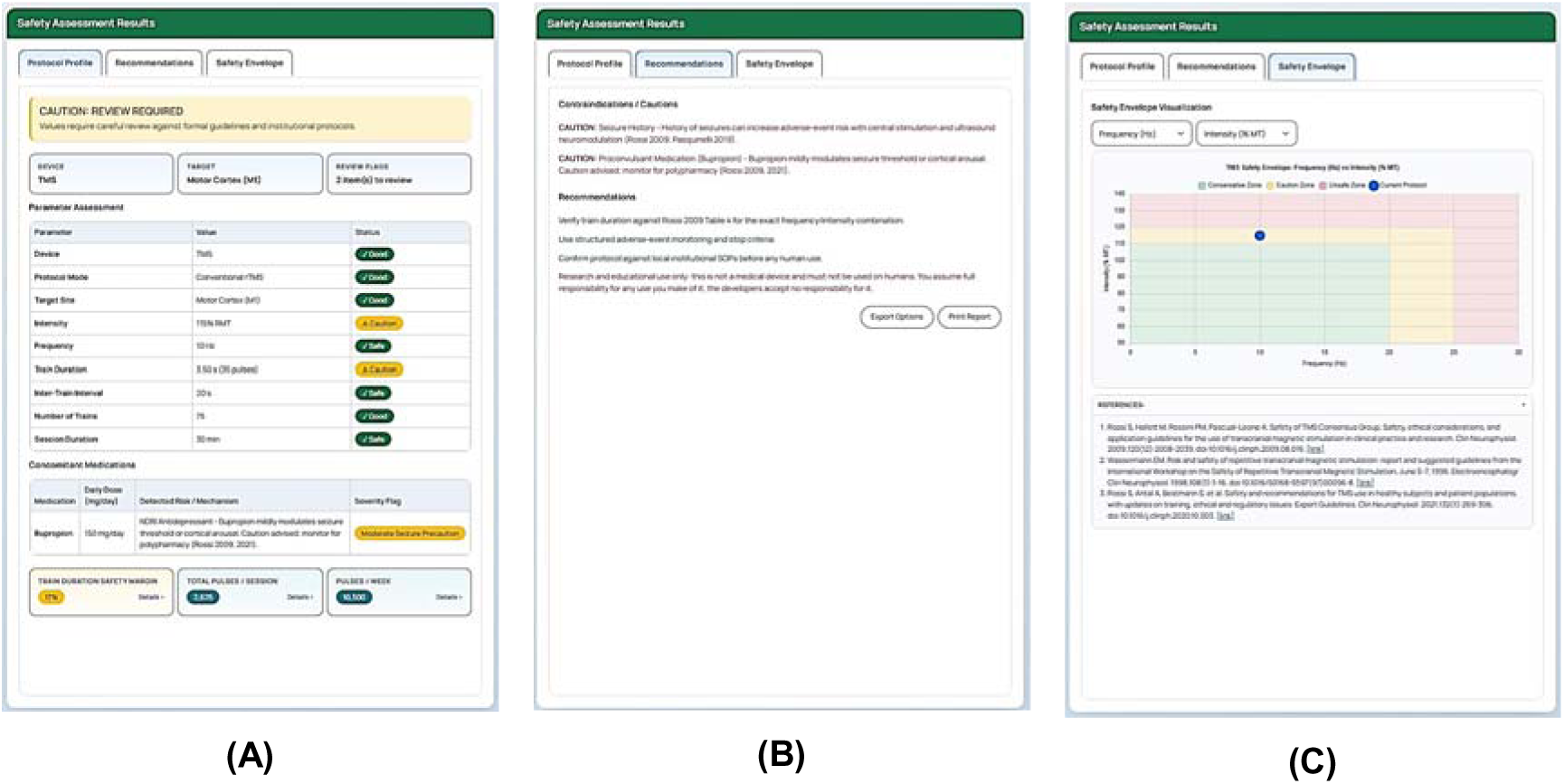
Safety Assessment Results panel of the graphical user interface, with its three tabs: (A) Protocol Profile: overall verdict with the per-parameter assessment table; (B) Recommendations: contraindication and caution findings, each carrying its threshold’s provenance; (C) Safety Envelope: two-dimensional view placing the submitted protocol relative to the modality’s boundary regions.

## Discussion

In this paper we presented the Explorer, a client-side web application that evaluates planned non-invasive neuromodulation protocols against published safety boundaries before laboratory documentation. Its defining property is that all 82 rules (29 literature-derived, 36 literature-anchored, 17 operational defaults) carry an explicit provenance classification across three transparent tiers. Where the literature supplies a trial parameter or adverse-event observation point rather than a consensus limit, the engine applies an explicit literature-anchored non-limit marker instead of attaching an unsubstantiated safety citation [16–20, 22]. As detailed in Section 3, the engine reproduced the declared verdict for all 38 example protocols spanning the six supported modalities (12 TMS, 5 tDCS, 5 PMS, 5 TENS, 5 tUS, 6 tVNS; declared bands SAFE 19, CAUTION 8, UNSAFE 11), with the static data and accessibility checks passing in the same run. The tool also exports protocol-level audit reports recording the specific provenance behind every verdict.

The scope of this evidence deserves plain statement. The verification suite is a *simulated* evaluation; it shows that the engine computes the verdicts its encoded rules require, and that the quality-assurance harness, including behavioural checks driven in a real browser and contrast measurements of rendered text styles, holds at the pinned version v1.1.0-beta. It does not constitute clinical validation, usability evaluation, or any assessment with human participants. The Explorer is a research and educational instrument bridging the safety-assurance gap in human laboratory protocols, not a medical device. It provides no clearance to stimulate a human subject. Responsibility for clinical risk management and any stimulation procedure remains strictly with the investigator, applicable consensus documents, screening procedures, and institutional oversight [15–17, 34].

From a regulatory perspective, medical device qualification across jurisdictions depends strictly on intended purpose rather than liability disclaimers. Under Brazilian Health Surveillance Agency regulations (ANVISA RDC No. 657/2022, Art. 4), European Union Medical Device Regulation (EU MDR 2017/745, Annex VIII, Rule 11), and US FDA Clinical Decision Support Software Guidance (2022), pre-investigational bench tools that automate parameter arithmetic verification without individualized clinical decision support are explicitly exempt from medical device registration. Accordingly, the “audit trails” generated by the Explorer serve strictly as reproducible experimental parameter logs for laboratory governance, not clinical trial compliance records.

Several design decisions behind the engine deserve explanation. The rule engine is deterministic rather than inferential. Consensus safety guidelines are themselves structured tabular boundaries [15–20], and an auditable implementation directly mirrors their structure, so that every deviation from a recommended threshold maps transparently to published evidence rather than heuristic extrapolation. Thresholds are also declared at a single point in the codebase, which genuinely prevents provenance decay and enables the verification suite to mechanically re-derive the safety envelope at every run. Just as importantly, the tool complements existing safety instruments rather than replacing them. Questionnaires remain the correct filter for subject-level contraindications [34]; field-modelling suites (such as SimNIBS and ROAST for electric/magnetic stimulation, or BabelBrain and k-Plan for acoustic simulation) solve the complementary problem of individual anatomical spatial dosimetry; and firmware interlocks enforce hardware ceilings, not consensus boundaries. Informal laboratory spreadsheets are precisely the operational error class the Explorer is designed to remove. Spanning six modalities in a unified interface supports the multi-modal protocols increasingly standard in contemporary laboratories, while modular isolation ensures breadth does not compromise modality-specific dosimetric depth. The pharmacological module likewise supplements, and does not replace, investigator judgement regarding medication effects [17, 32, 33]. Finally, as noted for focused ultrasound (tUS), consensus safety metrics [10] require in-situ acoustic exposure evaluation. Because the Explorer performs client-side parameter screening without patient-specific numerical simulation, skull derating must be computed prior to data entry to avoid artificially distorting exposure estimates.

This tier-aware verdict architecture addresses the operational tension between consensus safety ceilings and empirical clinical envelopes directly. In earlier iterations of the rule engine, empirical observation points (such as the 0.06 mA/cm² skin-lesion observation reported by Antal et al. [20]) or operational defaults operated as flat rejection barriers, producing false-positive alarms on mainstream protocols (such as 2.0 mA on 25 cm² tDCS, or SNT/SAINT accelerated rTMS [4, 5]). By explicitly segregating hard consensus limits (Tier 1) from empirical trial baselines (Tier 2) and operational precautions (Tier 3), the engine preserves uncompromising enforcement of physiological safety thresholds while providing nuanced specialist-review and pacing alerts for novel protocols. By evaluating continuous scalar thresholds through systematic boundary-value testing (*T* ± ɛ), the implementation verifies that tier transitions occur monotonically without edge anomalies, addressing finite coverage in discrete protocol libraries.

The limitations of the Explorer follow from what the tool is and what it is not. The rule set reflects consensus documents published at the time of encoding, and it will age as those guidelines are revised; each revision requires a deliberate re-encoding step, and the per-rule traceability makes such drift visible rather than preventing it. Coverage is bounded by the curated literature, so absent thresholds for rare parameter combinations trigger conservative defaults that stand in for missing evidence. The medication engine screens against documented sources [17, 32, 33] and cannot claim pharmacological completeness. The drug database covers 53 substances; the absence of an alert for an uncatalogued compound does not imply the absence of pharmacological risk. The application encodes safety boundaries without an efficacy engine, making no statements about treatment effect. Finally, the evaluation reported here is simulated and descriptive; no human usability study, no field deployment study, and no inter-rater comparison against expert screening has been conducted, so the tool’s fitness for routine laboratory workflow remains an open question.

Future work follows directly from these limitations. Priority items include an expert review of the encoded rules against consensus documents, a structured mechanism for tracking guideline updates into the engine, human usability evaluation of the screening workflow, and ultimately evidence from real-world laboratory documentation practices. Each of these would strengthen the evidence class of the evaluation, and each is precisely the reason this paper is careful to call its evaluation simulated rather than validated.

## Conclusion

We presented the Explorer, a browser-based tool that consolidates the safety thresholds scattered across the consensus literature for six non-invasive neuromodulation modalities into a single, source-provenanced rule engine. Every numeric boundary is categorized across three provenance tiers (literature-derived, literature-anchored, or operational default), so a user can see not only what the tool concluded but on what authority it concluded it. An automated internal verification suite of 38 example protocols, together with automated static integrity assertions, regression guards, and a behavioral and accessibility unit suite, demonstrated full engine-versus-declaration agreement, establishing the tool’s internal self-consistency under the version pinned for this paper (v1.1.0-beta).

The boundaries on interpretation bear repeating. The evaluation is simulated; it verifies the engine against its own declared rules rather than clinical accuracy, and no human evaluation has been performed. The tool’s intended use is research and education, so it is not a medical device, it is not cleared for clinical decision-making, and it must not be used to guide treatment in humans. The rule corpus, finally, is a snapshot that will require maintenance as the source guidelines are updated.

Within those limits, the tool offers a transparent, auditable substrate for parameter screening and safety reasoning in NINM research and teaching.

## Supporting information

supplementary-drug-database

supplementary-rule-inventory

supplementary-tests-results

## Data Availability

The Explorer is open-source under the MIT License and publicly hosted at https://github.com/Non-invasive-Neuromodulation-Lab/NINM_Safety (live deployment: https://neuromod.pesquisa.ufabc.edu.br/NINM_Safety/).

## Use of AI

The authors declare that they used GLM 5.3, DeepSeek 4.1-flash, and Gemini 3.8-flash for code structuring, testing harnesses, drafting assistance, text structuring, and language editing. However, all scientific content, safety threshold values, bibliographic entries, pharmacological interactions, and verification outcomes were derived from the authors’ own curated sources and stored run artifacts, and ultimate responsibility for the published content belongs exclusively to the authors. No AI tool was used for data analysis or image generation, and no AI tool is listed as an author.

## Authors’ contributions

Authors TSL idealized the contribution and evaluated the clinical aspects; YZ developed the code and drafted the MS; IRF curated the code. All authors approved the final version to be published and agreed to take public responsibility for all aspects of the study.

## Ethics statement

This work did not involve human participants or personal data. The tool evaluates simulated example protocols against published safety thresholds; no clinical data and no human evaluation were performed, so no research-ethics-committee approval or CAAE applies.

## Supplementary material

Supplementary Table S1. (full rule-by-rule inventory of the knowledge base, generated mechanically from the pinned build), Supplementary Table S2 (complete drug database: compounds, interaction classes, and evaluation basis), Supplementary File S3 (medical history screening specifications) and File S4 (tests results) accompany this paper.

