## Supplementary material for "NINM Safety Parameter Explorer: a safety-reference tool for non-invasive neuromodulation": Table 5

| **Element** | **Value / finding** |
| --- | --- |
| Inputs | tDCS, anodal, DLPFC; current 2.00 mA; electrode area 25 cm²; duration 30 min; 5 sessions/week |
| Current | mA — safe (below consensus conventional envelope 4 mA [17, 19]; empirical stroke escalation study [20] confirms tolerability at this boundary; common operational range 1–2 mA is an informational reference range, not a consensus safety limit) |
| Current density | $J=I/A=2.00/25.00=0.0800$ mA/cm² — caution (literature-anchored: published value, not a consensus safety limit; ceiling 0.057 mA/cm², adopted from published adverse-event observation [19]; specialist review advised) |
| Charge density | $Q/A=((2.00/1000)\times1800)/25.00=0.144$ C/cm² — safe (evaluated against Liebetanz et al. 2009 rat cortical lesion threshold of 5.24 C/cm² cited in Antal et al. [19]) |
| Total charge / session | C ($Q=(2.00/1000)\times1800$) — safe (within the $\leq$7.2 C conventional human trial session charge envelope [17]) |
| Session duration | min — safe ($\leq$40 min envelope [17]) |
| Weekly charge density | $Q_{\text{wk}}=5\times0.144=0.72$ C/cm²/wk — safe (conservative weekly limit 12 C/cm²/wk; unsourced operational default) |
| Overall verdict | CAUTION (outside 35 cm² conventional envelope; skin inspection advised; formerly evaluated as UNSAFE PARAMETER PROFILE) |
| Engine warning (verbatim) | “Current density (0.080 mA/cm²) exceeds the published observation point of $\leq$0.06 mA/cm², at which repeated daily tDCS ‘caused persisting skin lesions under the electrodes in some subjects’ (Antal 2017, at 25–35 cm² and 1.5–2.1 mA).” |
| Operational default (verbatim labelling for weekly charge cap) | “UNSOURCED — this is an operational default, not literature-derived: no consensus guideline specifies this value, and it is retained as a precaution rather than presented as cited.” |
