## supplementary-drug-database for "NINM Safety Parameter Explorer: a safety-reference tool for non-invasive neuromodulation"

### **Supplementary Material — Table S2. Medication database of the NINM Safety Explorer**

#### **Scope**

The screening engine considers **only** the 53 substances listed below (225 recognised name strings, 225 unique). Any medication string that resolves to none of these entries receives the tool’s **Unmonitored / Uncatalogued** descriptor: no pharmacological profile, no interaction evaluation, and no pharmacological warning is produced for it. Standard references cited in the database header: Rossi 2009; Rossi 2021; Antal 2017; Ziemann 2015; McLaren 2018. Per-substance provenance, including conservative-default and external-regulatory-source attributions, are reproduced verbatim in each entry’s Provenance line.

#### **Summary**

| **Measure** | **Count** |
| --- | --- |
| Catalogued substances | 53 |
| Recognised name strings (aliases incl. canonical) | 225 |
| Unique name strings | 225 |
| isHighSeizureRisk = true | 18 |
| isPlasticityModifier = true | 19 |
| isProconvulsant = true | 37 |
| isSeizureRisk = true | 38 |
| isStimulant = true | 5 |
| isTca = true | 6 |
| Entries with alert-override text | 12 |

#### **Full inventory**

##### **Clozapine (clozapine)**

- **Class:** Atypical Antipsychotic
- **Recognised names:** clozapine, clozapina, leponex, fazaclo, versacloz
- **Flags (true):** isHighSeizureRisk, isSeizureRisk, isProconvulsant
- **Provenance (verbatim mechanism field):** Strong proconvulsant; markedly lowers seizure threshold in a dose-dependent manner; named in Rossi 2009 §5.3 List 1 and Rossi 2021 as a high seizure hazard.

##### **Chlorpromazine (chlorpromazine)**

- **Class:** Phenothiazine Antipsychotic
- **Recognised names:** chlorpromazine, clorpromazina, amplictil, thorazine, largactil
- **Flags (true):** isHighSeizureRisk, isSeizureRisk, isProconvulsant
- **Provenance (verbatim mechanism field):** Dopamine antagonist; high proconvulsant potential among typical antipsychotics; named in Rossi 2009 §5.3 List 1 and Rossi 2021 as a high seizure hazard.

##### **Maprotiline (maprotiline)**

- **Class:** Tetracyclic Antidepressant
- **Recognised names:** maprotiline, maprotilina, ludiomil
- **Flags (true):** isHighSeizureRisk, isSeizureRisk, isProconvulsant
- **Provenance (verbatim mechanism field):** High epileptogenic potential; significantly lowers seizure threshold; named in Rossi 2009 §5.3 List 1 and Rossi 2021 as a high seizure hazard.

##### **Doxepin (doxepin)**

- **Class:** Tricyclic Antidepressant (TCA)
- **Recognised names:** doxepin, doxepina, sinequan, zonalon, prudoxin, silenor
- **Flags (true):** isHighSeizureRisk, isSeizureRisk, isProconvulsant, isTca
- **Alert override:** yes (engine uses an entry-specific warning text)
- **Provenance (verbatim mechanism field):** Tricyclic antidepressant with potent seizure-threshold lowering potential; named in Rossi 2009 §5.3 List 1 as a strong proconvulsant hazard.

##### **Cocaine (cocaine)**

- **Class:** CNS Stimulant / Local Anesthetic (Substance of Abuse)
- **Recognised names:** cocaine, cocaína, cocaina
- **Flags (true):** isHighSeizureRisk, isSeizureRisk, isProconvulsant, isStimulant
- **Alert override:** yes (engine uses an entry-specific warning text)
- **Provenance (verbatim mechanism field):** Central nervous system stimulant and local anesthetic; named in Rossi 2009 §5.3 List 1 as a strong proconvulsant hazard that significantly lowers seizure threshold.

##### **MDMA (Ecstasy) (mdma)**

- **Class:** Amphetamine Derivative / Entactogen
- **Recognised names:** mdma, ecstasy, êxtase, extase
- **Flags (true):** isHighSeizureRisk, isSeizureRisk, isProconvulsant, isStimulant
- **Alert override:** yes (engine uses an entry-specific warning text)
- **Provenance (verbatim mechanism field):** Substituted amphetamine with central monoaminergic release; named in Rossi 2009 §5.3 List 1 as a strong hazard with high proconvulsant potential.

##### **Phencyclidine (PCP) (pcp)**

- **Class:** Dissociative Anesthetic / Hallucinogen
- **Recognised names:** phencyclidine, pcp, fenciclidina
- **Flags (true):** isHighSeizureRisk, isSeizureRisk, isProconvulsant, isPlasticityModifier
- **Plasticity type:** nmda_antagonist
- **Alert override:** yes (engine uses an entry-specific warning text)
- **Provenance (verbatim mechanism field):** Dissociative NMDA receptor antagonist; named in Rossi 2009 §5.3 List 1 as a strong hazard with substantial seizure-threshold lowering potential.

##### **Gamma-Hydroxybutyrate (GHB) (ghb)**

- **Class:** CNS Depressant / GABA-B Agonist
- **Recognised names:** ghb, gamma-hydroxybutyrate, gamma-hidroxibutirato, sodium oxybate, oxibato de sódio, xyrem
- **Flags (true):** isHighSeizureRisk, isSeizureRisk, isProconvulsant
- **Alert override:** yes (engine uses an entry-specific warning text)
- **Provenance (verbatim mechanism field):** GABA-B and GHB receptor agonist; named in Rossi 2009 §5.3 List 1 as a strong proconvulsant hazard that produces spike-wave discharges and lowers seizure threshold.

##### **Foscarnet (foscarnet)**

- **Class:** Antiviral Agent
- **Recognised names:** foscarnet, foscarneto, foscavir
- **Flags (true):** isHighSeizureRisk, isSeizureRisk, isProconvulsant
- **Alert override:** yes (engine uses an entry-specific warning text)
- **Provenance (verbatim mechanism field):** Pyrophosphate analogue antiviral; named in Rossi 2009 §5.3 List 1 as a strong hazard due to mineral electrolyte chelation and high proconvulsant potential.

##### **Ganciclovir (ganciclovir)**

- **Class:** Antiviral Agent
- **Recognised names:** ganciclovir, ganciclovir sódico, cymevene, cytovene
- **Flags (true):** isHighSeizureRisk, isSeizureRisk, isProconvulsant
- **Alert override:** yes (engine uses an entry-specific warning text)
- **Provenance (verbatim mechanism field):** Nucleoside analogue antiviral; named in Rossi 2009 §5.3 List 1 as an antiviral agent with strong seizure-threshold lowering potential.

##### **Ritonavir (ritonavir)**

- **Class:** Antiviral / Protease Inhibitor
- **Recognised names:** ritonavir, norvir, kaletra
- **Flags (true):** isHighSeizureRisk, isSeizureRisk, isProconvulsant
- **Alert override:** yes (engine uses an entry-specific warning text)
- **Provenance (verbatim mechanism field):** HIV protease inhibitor and potent CYP3A4 inhibitor; named in Rossi 2009 §5.3 List 1 as an agent with strong seizure-threshold lowering potential and pharmacokinetic drug-interaction risks.

##### **Bupropion (bupropion)**

- **Class:** NDRI Antidepressant
- **Recognised names:** bupropion, bupropiona, bup, wellbutrin, zyban, zetron, bupium
- **Flags (true):** isHighSeizureRisk, isSeizureRisk, isProconvulsant
- **Provenance (verbatim mechanism field):** Dose-dependent proconvulsant risk; regulatory labeling reports dose-related seizure incidence (FDA bupropion prescribing information — external source, dailymed.nlm.nih.gov). The >= 300 mg/day escalation trigger is this app’s conservative default, not a published threshold.

##### **Amitriptyline (amitriptyline)**

- **Class:** Tricyclic Antidepressant (TCA)
- **Recognised names:** amitriptyline, amitriptilina, tryptanol, elavil, amytril
- **Flags (true):** isHighSeizureRisk, isSeizureRisk, isProconvulsant, isTca
- **Provenance (verbatim mechanism field):** Lowers seizure threshold; the > 100 mg/day escalation trigger is this app’s conservative default (Rossi 2009 §5.3 lists TCAs as a relative hazard without dose thresholds).

##### **Imipramine (imipramine)**

- **Class:** Tricyclic Antidepressant (TCA)
- **Recognised names:** imipramine, imipramina, tofranil
- **Flags (true):** isHighSeizureRisk, isSeizureRisk, isProconvulsant, isTca
- **Provenance (verbatim mechanism field):** Lowers seizure threshold; the > 100 mg/day escalation trigger is this app’s conservative default (Rossi 2009 §5.3 lists TCAs as a relative hazard without dose thresholds).

##### **Clomipramine (clomipramine)**

- **Class:** Tricyclic Antidepressant (TCA)
- **Recognised names:** clomipramine, clomipramina, anafranil
- **Flags (true):** isHighSeizureRisk, isSeizureRisk, isProconvulsant, isTca
- **Provenance (verbatim mechanism field):** Lowers seizure threshold; the > 100 mg/day escalation trigger is this app’s conservative default (Rossi 2009 §5.3 lists TCAs as a relative hazard without dose thresholds).

##### **Nortriptyline (nortriptyline)**

- **Class:** Tricyclic Antidepressant (TCA)
- **Recognised names:** nortriptyline, nortriptilina, pamelor
- **Flags (true):** isHighSeizureRisk, isSeizureRisk, isProconvulsant, isTca
- **Provenance (verbatim mechanism field):** Lowers seizure threshold; TCA proconvulsant caution applies (Rossi 2009 §5.3 List 2). The > 100 mg/day escalation trigger is this app’s conservative default.

##### **Desipramine (desipramine)**

- **Class:** Tricyclic Antidepressant (TCA)
- **Recognised names:** desipramine, desipramina, norpramin, pertofrane
- **Flags (true):** isHighSeizureRisk, isSeizureRisk, isProconvulsant, isTca
- **Provenance (verbatim mechanism field):** Lowers seizure threshold; the > 100 mg/day escalation trigger is this app’s conservative default (Rossi 2009 §5.3 lists TCAs as a relative hazard without dose thresholds).

##### **Sertraline (sertraline)**

- **Class:** SSRI Antidepressant
- **Recognised names:** sertraline, sertralina, zoloft, assert, serenata, tolrest
- **Flags (true):** isSeizureRisk, isProconvulsant
- **Provenance (verbatim mechanism field):** Selective serotonin reuptake inhibitor; mild seizure threshold reduction; caution with polypharmacy (Rossi 2009 §5.3 List 2; Rossi 2021).

##### **Fluoxetine (fluoxetine)**

- **Class:** SSRI Antidepressant
- **Recognised names:** fluoxetine, fluoxetina, prozac, daforin, fluxene, verotina
- **Flags (true):** isSeizureRisk, isProconvulsant
- **Provenance (verbatim mechanism field):** Selective serotonin reuptake inhibitor; mild seizure threshold reduction (Rossi 2009 §5.3 List 2; Rossi 2021).

##### **Escitalopram (escitalopram)**

- **Class:** SSRI Antidepressant
- **Recognised names:** escitalopram, lexapro, exodus, reconter
- **Flags (true):** isSeizureRisk, isProconvulsant
- **Provenance (verbatim mechanism field):** Selective serotonin reuptake inhibitor; mild seizure threshold reduction (Rossi 2009 §5.3 List 2; Rossi 2021).

##### **Paroxetine (paroxetine)**

- **Class:** SSRI Antidepressant
- **Recognised names:** paroxetine, paroxetina, paxil, pondera, aropax
- **Flags (true):** isSeizureRisk, isProconvulsant
- **Provenance (verbatim mechanism field):** Selective serotonin reuptake inhibitor; mild seizure threshold reduction (Rossi 2009 §5.3 List 2; Rossi 2021).

##### **Citalopram (citalopram)**

- **Class:** SSRI Antidepressant
- **Recognised names:** citalopram, celexa, cipramil, cipram, procimax, alcytam
- **Flags (true):** isSeizureRisk, isProconvulsant, isPlasticityModifier
- **Plasticity type:** ssri
- **Provenance (verbatim mechanism field):** Selective serotonin reuptake inhibitor; named in Rossi 2009 §5.3 List 2 as having potential to lower seizure threshold; McLaren 2018 notes citalopram enhances and consolidates tDCS-induced neuroplasticity.

##### **Venlafaxine (venlafaxine)**

- **Class:** SNRI Antidepressant
- **Recognised names:** venlafaxine, venlafaxina, effexor, efexor, alenthus
- **Flags (true):** isSeizureRisk, isProconvulsant
- **Provenance (verbatim mechanism field):** Serotonin-norepinephrine reuptake inhibitor; mild proconvulsant risk (Rossi 2009 §5.3 List 2; Rossi 2021).

##### **Duloxetine (duloxetine)**

- **Class:** SNRI Antidepressant
- **Recognised names:** duloxetine, duloxetina, cymbalta, velija, dual
- **Flags (true):** isSeizureRisk, isProconvulsant
- **Provenance (verbatim mechanism field):** Serotonin-norepinephrine reuptake inhibitor; mild proconvulsant risk (Rossi 2009 §5.3 List 2; Rossi 2021).

##### **Mirtazapine (mirtazapine)**

- **Class:** NaSSA Antidepressant
- **Recognised names:** mirtazapine, mirtazapina, remeron, menelat, razapina
- **Flags (true):** isSeizureRisk, isProconvulsant
- **Provenance (verbatim mechanism field):** Noradrenergic and specific serotonergic antidepressant; named in Rossi 2009 §5.3 List 2 as having potential to lower seizure threshold.

##### **Methylphenidate (methylphenidate)**

- **Class:** CNS Stimulant
- **Recognised names:** methylphenidate, metilfenidato, ritalin, ritalina, concerta, ragione
- **Flags (true):** isSeizureRisk, isProconvulsant, isStimulant
- **Provenance (verbatim mechanism field):** Dopamine-norepinephrine reuptake inhibitor; elevates central arousal; proconvulsant synergy risk (Rossi 2009 §5.3; Rossi 2021; Ziemann 2015).

##### **Lisdexamfetamine (lisdexamfetamine)**

- **Class:** CNS Stimulant
- **Recognised names:** lisdexamfetamine, lisdexanfetamina, vyvanse, venvanse, juneve
- **Flags (true):** isSeizureRisk, isProconvulsant, isStimulant
- **Provenance (verbatim mechanism field):** Dextroamphetamine prodrug; central sympathomimetic; proconvulsant synergy risk (Rossi 2009 §5.3; Rossi 2021).

##### **Dextroamphetamine / Amphetamine (dextroamphetamine)**

- **Class:** CNS Stimulant
- **Recognised names:** dextroamphetamine, amphetamine, anfetamina, adderall, dexedrine
- **Flags (true):** isSeizureRisk, isProconvulsant, isStimulant
- **Provenance (verbatim mechanism field):** Central nervous system stimulant; elevates arousal and proconvulsant synergy risk (Rossi 2009 §5.3; Rossi 2021; Ziemann 2015).

##### **Haloperidol (haloperidol)**

- **Class:** Typical Antipsychotic
- **Recognised names:** haloperidol, haldol
- **Flags (true):** isSeizureRisk, isProconvulsant
- **Provenance (verbatim mechanism field):** Dopamine D2 receptor antagonist; moderate seizure threshold reduction (Rossi 2009 §5.3 List 2; Rossi 2021; Ziemann 2015).

##### **Olanzapine (olanzapine)**

- **Class:** Atypical Antipsychotic
- **Recognised names:** olanzapine, olanzapina, zyprexa
- **Flags (true):** isSeizureRisk, isProconvulsant
- **Provenance (verbatim mechanism field):** Second-generation antipsychotic; moderate seizure threshold reduction (Rossi 2009 §5.3 List 2; Rossi 2021).

##### **Quetiapine (quetiapine)**

- **Class:** Atypical Antipsychotic
- **Recognised names:** quetiapine, quetiapina, seroquel
- **Flags (true):** isSeizureRisk, isProconvulsant
- **Provenance (verbatim mechanism field):** Second-generation antipsychotic; moderate seizure threshold reduction (Rossi 2009 §5.3 List 2; Rossi 2021).

##### **Risperidone (risperidone)**

- **Class:** Atypical Antipsychotic
- **Recognised names:** risperidone, risperidona, risperdal
- **Flags (true):** isSeizureRisk, isProconvulsant
- **Provenance (verbatim mechanism field):** Second-generation antipsychotic; moderate seizure threshold reduction (Rossi 2009 §5.3 List 2; Rossi 2021).

##### **Aripiprazole (aripiprazole)**

- **Class:** Atypical Antipsychotic
- **Recognised names:** aripiprazole, aripiprazol, abilify, aristab
- **Flags (true):** isSeizureRisk, isProconvulsant
- **Provenance (verbatim mechanism field):** Partial dopamine D2 and 5-HT1A agonist; named in Rossi 2009 §5.3 List 2 as an antipsychotic with moderate proconvulsant potential.

##### **Lithium (lithium)**

- **Class:** Mood Stabilizer
- **Recognised names:** lithium, lítio, litio, carbolitium, eskalith, lithobid
- **Flags (true):** isSeizureRisk, isProconvulsant, isPlasticityModifier
- **Plasticity type:** mood_stabilizer
- **Provenance (verbatim mechanism field):** Mood stabilizer; named in Rossi 2009 §5.3 List 2 as carrying relative proconvulsant potential; modulates intracellular signaling cascades and neuroplasticity (Ziemann 2015).

##### **Carbamazepine (carbamazepine)**

- **Class:** Sodium Channel Blocker / Anticonvulsant
- **Recognised names:** carbamazepine, carbamazepina, tegretol
- **Flags (true):** isPlasticityModifier
- **Plasticity type:** channel_blocker
- **Provenance (verbatim mechanism field):** Blocks voltage-gated sodium channels; elevates motor threshold and suppresses tDCS neuroplasticity (Ziemann 2015).

##### **Lamotrigine (lamotrigine)**

- **Class:** Sodium/Calcium Channel Blocker / Anticonvulsant
- **Recognised names:** lamotrigine, lamotrigina, lamictal, neural
- **Flags (true):** isPlasticityModifier
- **Plasticity type:** channel_blocker
- **Provenance (verbatim mechanism field):** Inhibits voltage-gated Na+/Ca2+ channels; elevates motor threshold and reduces PAS-induced LTP-like plasticity (Ziemann 2015). A tDCS-specific interaction is not established in held sources.

##### **Phenytoin (phenytoin)**

- **Class:** Sodium Channel Blocker / Anticonvulsant
- **Recognised names:** phenytoin, fenitoina, fenitoína, dilantin, hidantal
- **Flags (true):** isPlasticityModifier
- **Plasticity type:** channel_blocker
- **Provenance (verbatim mechanism field):** Voltage-gated sodium channel blocker; reduces cortical excitability and blocks tDCS plasticity (Ziemann 2015).

##### **Oxcarbazepine (oxcarbazepine)**

- **Class:** Voltage-Gated Ion Channel Blocker / Anticonvulsant
- **Recognised names:** oxcarbazepine, oxcarbazepina, trileptal
- **Flags (true):** isPlasticityModifier
- **Plasticity type:** channel_blocker
- **Provenance (verbatim mechanism field):** Modulates voltage-sensitive Na+/Ca2+ channels; dampens cortical excitability and plasticity (Ziemann 2015; Antal 2017).

##### **Valproic Acid / Divalproex (valproate)**

- **Class:** GABA Enhancer / HDAC Inhibitor / Anticonvulsant
- **Recognised names:** valproate, valproic acid, acido valproico, ácido valproico, divalproex, depakene, depakote, torval
- **Flags (true):** isPlasticityModifier
- **Plasticity type:** gaba_enhancer_hdac_inhibitor
- **Provenance (verbatim mechanism field):** Broad-spectrum anticonvulsant; enhances GABAergic transmission, inhibits histone deacetylases (HDAC), and attenuates sodium channel conductance; elevates motor threshold and modulates neuroplasticity (Ziemann 2015; Rossi 2009 §5.3).

##### **Memantine (memantine)**

- **Class:** NMDA Receptor Antagonist
- **Recognised names:** memantine, memantina, ebix, namenda, alois
- **Flags (true):** isPlasticityModifier
- **Plasticity type:** nmda_antagonist
- **Provenance (verbatim mechanism field):** Uncompetitive NMDA receptor antagonist; abolishes LTP-like plasticity induced by iTBS and laser-PAS (Ziemann 2015). A tDCS-specific effect is not established in held sources.

##### **Dextromethorphan (dextromethorphan)**

- **Class:** NMDA Receptor Antagonist / Antitussive
- **Recognised names:** dextromethorphan, dextrometorfano, silomat, benylin
- **Flags (true):** isPlasticityModifier
- **Plasticity type:** nmda_antagonist
- **Provenance (verbatim mechanism field):** Non-competitive NMDA receptor blocker; suppresses LTP-like plasticity consolidation in tDCS (McLaren 2018; Ziemann 2015).

##### **Ketamine (ketamine)**

- **Class:** NMDA Receptor Antagonist / Dissociative Anesthetic
- **Recognised names:** ketamine, cetamina, ketalar
- **Flags (true):** isSeizureRisk, isPlasticityModifier
- **Plasticity type:** nmda_antagonist
- **Alert override:** yes (engine uses an entry-specific warning text)
- **Provenance (verbatim mechanism field):** Potent NMDA receptor blocker; profoundly alters cortical plasticity and excitability. Named in Rossi 2009 §5.3 list 1 (significant seizure-threshold lowering potential); Ziemann 2015 reports dose-dependent motor-threshold decrease (increased corticospinal excitability).

##### **Clonazepam (clonazepam)**

- **Class:** Benzodiazepine (GABA-A Allosteric Modulator)
- **Recognised names:** clonazepam, rivotril, klonopin, clonavita
- **Flags (true):** isPlasticityModifier
- **Plasticity type:** gaba_modulator
- **Provenance (verbatim mechanism field):** Positive allosteric modulator of GABA-A receptors; increases SICI and MEP-amplitude effects on TMS measures (Ziemann 2015). Acute withdrawal or abrupt cessation forms a strong seizure hazard (Rossi 2009 §5.3 List 3). tDCS evidence is mixed: lorazepam initially delayed then prolonged anodal after-effects physiologically (Nitsche 2004 via McLaren 2018), while Antal 2017 cautions benzodiazepines may reduce clinical efficacy.

##### **Diazepam (diazepam)**

- **Class:** Benzodiazepine (GABA-A Allosteric Modulator)
- **Recognised names:** diazepam, valium, valbra, dienpax
- **Flags (true):** isPlasticityModifier
- **Plasticity type:** gaba_modulator
- **Provenance (verbatim mechanism field):** Enhances GABAergic inhibition; increases motor threshold and SICI on TMS measures (Ziemann 2015). Acute withdrawal or abrupt cessation forms a strong seizure hazard (Rossi 2009 §5.3 List 3). tDCS plasticity evidence is mixed in held sources (see lorazepam, Nitsche 2004 via McLaren 2018).

##### **Lorazepam (lorazepam)**

- **Class:** Benzodiazepine (GABA-A Allosteric Modulator)
- **Recognised names:** lorazepam, ativan, lorax
- **Flags (true):** isPlasticityModifier
- **Plasticity type:** gaba_modulator
- **Provenance (verbatim mechanism field):** Potentiates GABA-A signaling; increases SICI on TMS measures (Ziemann 2015). Acute withdrawal or abrupt cessation forms a strong seizure hazard (Rossi 2009 §5.3 List 3). tDCS evidence is mixed: lorazepam initially delayed then prolonged anodal after-effects (Nitsche 2004 via McLaren 2018).

##### **Alprazolam (alprazolam)**

- **Class:** Benzodiazepine (GABA-A Allosteric Modulator)
- **Recognised names:** alprazolam, frontal, xanax, apraz
- **Flags (true):** isPlasticityModifier
- **Plasticity type:** gaba_modulator
- **Provenance (verbatim mechanism field):** Positive allosteric modulator of GABA-A; increases SICI on TMS measures (Ziemann 2015). Acute withdrawal or abrupt cessation forms a strong seizure hazard (Rossi 2009 §5.3 List 3). tDCS-specific plasticity effects are mixed in held sources.

##### **Zolpidem (zolpidem)**

- **Class:** Non-benzodiazepine GABA-A Agonist (Z-drug)
- **Recognised names:** zolpidem, stilnox, ambien, patz, noctiden
- **Flags (true):** isPlasticityModifier
- **Plasticity type:** gaba_modulator
- **Provenance (verbatim mechanism field):** Selective alpha-1 GABA-A receptor agonist; alters cortical excitability and sleep architecture (Ziemann 2015; Antal 2017).

##### **Phenobarbital (phenobarbital)**

- **Class:** Barbiturate (GABA-A Agonist / Channel Modulator)
- **Recognised names:** phenobarbital, fenobarbital, gardenal, luminal
- **Flags (true):** isPlasticityModifier
- **Plasticity type:** gaba_modulator
- **Provenance (verbatim mechanism field):** Enhances GABA-mediated chloride flux and blocks AMPA channels; elevates motor threshold (Ziemann 2015). Acute withdrawal or abrupt cessation forms a strong seizure hazard (Rossi 2009 §5.3 List 3).

##### **Alcohol (alcohol)**

- **Class:** CNS Depressant (Ethanol)
- **Recognised names:** alcohol, ethanol, álcool, etanol
- **Flags (true):** isSeizureRisk, isProconvulsant
- **Alert override:** yes (engine uses an entry-specific warning text)
- **Provenance (verbatim mechanism field):** Acute intoxication and, especially, withdrawal lower the seizure threshold: Rossi 2009 §5.3 places alcohol on the strong-hazard list and names its withdrawal a strong relative hazard for rTMS; Rossi 2021 names alcohol consumption among seizure-threshold-lowering factors and recommends documenting it. Note: the dedicated screening path for the withdrawal/intoxication state is the medical-history checkbox (relative_high + specialist review for TMS/tDCS); this entry lets free-text medication entry also resolve alcohol. Generic names only (INN + Portuguese); no dose threshold exists in the held sources.

##### **Theophylline (theophylline)**

- **Class:** Methylxanthine / Phosphodiesterase Inhibitor
- **Recognised names:** theophylline, teofilina
- **Flags (true):** isHighSeizureRisk, isSeizureRisk, isProconvulsant
- **Alert override:** yes (engine uses an entry-specific warning text)
- **Provenance (verbatim mechanism field):** Named verbatim in Rossi 2009 §5.3 list 1 (“significant seizure threshold lowering potential”); the guideline calls it arguably the classic drug-seizure interaction example. No dose threshold is stated in the held sources.

##### **Tramadol (tramadol)**

- **Class:** Opioid Analgesic
- **Recognised names:** tramadol, tramadol hydrochloride, cloridrato de tramadol
- **Flags (true):** isSeizureRisk, isProconvulsant
- **Alert override:** yes (engine uses an entry-specific warning text)
- **Provenance (verbatim mechanism field):** Well-documented seizure-threshold-lowering analgesic per regulatory labeling: seizure risk is a labeled warning in the FDA tramadol prescribing information (external regulatory source, dailymed.nlm.nih.gov). No dose threshold is stated in the held sources, so none is applied.

##### **Levodopa (L-DOPA) (levodopa)**

- **Class:** Dopamine Precursor / Antiparkinson Agent
- **Recognised names:** levodopa, l-dopa, prolopa, sinemet
- **Flags (true):** isPlasticityModifier
- **Plasticity type:** dopamine_agonist
- **Provenance (verbatim mechanism field):** Dopamine precursor; modulates cortical neuroplasticity, prolonging and enhancing tDCS after-effects in a dosage-dependent manner (McLaren 2018; Ziemann 2015).

##### **Rivastigmine (rivastigmine)**

- **Class:** Cholinesterase Inhibitor
- **Recognised names:** rivastigmine, rivastigmina, exelon, prometax
- **Flags (true):** isPlasticityModifier
- **Plasticity type:** cholinergic_enhancer
- **Provenance (verbatim mechanism field):** Acetylcholinesterase inhibitor; restores and enhances impaired tDCS-induced neuroplasticity in elderly and neurodegenerative populations (McLaren 2018).
