## supplementary-rule-inventory for "NINM Safety Parameter Explorer: a safety-reference tool for non-invasive neuromodulation"

### **Supplementary Material — Table S1. Full rule inventory of the NINM Safety Explorer**

#### **Summary**

| **Class** | **Count** |
| --- | --- |
| Modality safety rules | 82 |
| — Literature-derived (consensus guidelines / primary literature) | 11 |
| — Literature-anchored (published clinical values, NOT consensus safety limits) | 54 |
| — Operational defaults (precautionary engineering baselines) | 17 |
| Target-site session-duration caps (TMS sites; 4 anchored, 1 default) | 5 |
| Total evaluative safety rules | 87 |
| Informational reference values (advisory context, non-evaluative) | 15 |
| Target-site display-name labels (not rules) | 21 |
| Citation metadata strings (not rules) | 2 |
| Rossi 2009 Table 4 train-duration maxima (special table) | 5 frequency rows $\times$ 5 intensities = 25 cells |
| Rossi Table 4 "$>$" tested-maxima markings (special set) | 13 |
| TBS protocol definitions (Huang 2005; special table) | 2 |
| Citation registry (SAFETY_REFERENCES) | 36 entries in 6 modality groups (36 unique ids) |

#### **Per modality**

| **Modality** | **Literature-derived** | **Literature-anchored** | **Operational default** | **Total Rules** | **Informational** |
| --- | --- | --- | --- | --- | --- |
| TMS | 1 | 10 | 4 | 15 | 0 |
| tDCS | 3 | 5 | 3 | 11 | 5 |
| PMS | 0 | 4 | 4 | 8 | 0 |
| TENS | 0 | 6 | 1 | 7 | 0 |
| tUS | 7 | 13 | 3 | 23 | 2 |
| tVNS | 0 | 16 | 2 | 18 | 8 |
| **All** | **11** | **54** | **17** | **82** | **15** |

TBS has no threshold-tree leaves: its two fixed protocol definitions are special-case tables (Huang 2005), listed in the Summary above.

*Note on variable nomenclature and regulatory benchmarks:* Variable names in the inventory reflect the standardized implementation identifiers in v1.1.0-beta (tus.pulseDurationSafeMaxMs = 5 ms, tus.pulseDurationCautionMaxMs = 10 ms), where values up to 5 ms are classified as safe, exposures from 5 to 10 ms trigger caution, and exposures exceeding 10 ms trigger an unsafe verdict. Published regulatory reference values (such as the FDA diagnostic ultrasound $I_{\text{SPPA}}$ limit of 190 W/cm^2^) are categorized as non-evaluative informational reference values providing advisory context without altering safety verdicts.

#### **Full inventory**

| **Path** | **Value** | **Unit** | **Tier** | **Effect on Verdict** | **Citation + Locator** | **Verbatim Quote** |
| --- | --- | --- | --- | --- | --- | --- |
| targetSites.motor_cortex.name | "Motor Cortex (M1)" | — | informational | None (display label) | None (display label) | Target site display label. |
| targetSites.motor_cortex.tms.maxSessionDuration |  | min | literature-anchored | $>$60 min triggers caution verdict | Rossi 2009, § 3.2; Lefaucheur 2020, § 3.1 | Motor cortex rTMS session durations in published literature typically $\leq$60 min. |
| targetSites.dlpfc.name | "Dorsolateral Prefrontal Cortex (DLPFC)" | — | informational | None (display label) | None (display label) | Target site display label. |
| targetSites.dlpfc.tms.maxSessionDuration |  | min | literature-anchored | $>$40 min triggers caution verdict | Rossi 2009, § 3.2; Lefaucheur 2020, § 3.1 | DLPFC depression rTMS protocols typically $\leq$40 min (Rossi 2009, Lefaucheur 2020). |
| targetSites.cingulate.name | "Anterior Cingulate Cortex (ACC)" | — | informational | None (display label) | None (display label) | Target site display label. |
| targetSites.cingulate.tms.maxSessionDuration |  | min | literature-anchored | $>$40 min triggers caution verdict | Rossi 2009, § 3.2; Lefaucheur 2020, § 3.1 | Anterior cingulate cortex stimulation protocols typically $\leq$40 min. |
| targetSites.visual.name | "Visual Cortex" | — | informational | None (display label) | None (display label) | Target site display label. |
| targetSites.visual.tms.maxSessionDuration |  | min | literature-anchored | $>$30 min triggers caution verdict | Rossi 2009, § 3.2 | Visual cortex phosphene/excitability protocols typically $\leq$30 min. |
| targetSites.upper_limb.name | "Upper Limb" | — | informational | None (display label) | None (display label) | Target site display label. |
| targetSites.lower_limb.name | "Lower Limb" | — | informational | None (display label) | None (display label) | Target site display label. |
| targetSites.pelvic.name | "Pelvic Floor / Sacral Region" | — | informational | None (display label) | None (display label) | Target site display label. |
| targetSites.lumbar.name | "Lumbar / Back" | — | informational | None (display label) | None (display label) | Target site display label. |
| targetSites.cervical.name | "Cervical / Neck" | — | informational | None (display label) | None (display label) | Target site display label. |
| targetSites.thalamus.name | "Thalamus" | — | informational | None (display label) | None (display label) | Target site display label. |
| targetSites.cervical_spine.name | "Cervical Spinal Column (C2–T1)" | — | informational | None (display label) | None (display label) | Target site display label. |
| targetSites.thoracic_spine.name | "Thoracic Spinal Column (T1–T12)" | — | informational | None (display label) | None (display label) | Target site display label. |
| targetSites.lumbar_spine.name | "Lumbar Spinal Column (L1–L5)" | — | informational | None (display label) | None (display label) | Target site display label. |
| targetSites.sacral_spine.name | "Sacral Region" | — | informational | None (display label) | None (display label) | Target site display label. |
| targetSites.auricular_vns.name | "Auricular (Ear) Vagus Branch" | — | informational | None (display label) | None (display label) | Target site display label. |
| targetSites.cervical_vns.name | "Cervical Vagus Nerve" | — | informational | None (display label) | None (display label) | Target site display label. |
| targetSites.hippocampus.name | "Hippocampus" | — | informational | None (display label) | None (display label) | Target site display label. |
| targetSites.amygdala.name | "Amygdala" | — | informational | None (display label) | None (display label) | Target site display label. |
| targetSites.parietal.name | "Parietal Cortex" | — | informational | None (display label) | None (display label) | Target site display label. |
| targetSites.temporal.name | "Temporal Cortex" | — | informational | None (display label) | None (display label) | Target site display label. |
| targetSites.other.name | "Other / Unlabeled Site" | — | informational | None (display label) | None (display label) | Target site display label. |
| targetSites.other.tms.maxSessionDuration |  | min | operational default | $>$20 min triggers caution verdict | None (operational default) | Conservative default for unspecified target sites; no held guideline states a duration. |
| tms.maxPulsesPerSession |  | pulses | operational default | $>$6,000 pulses/session triggers caution review | None (operational default; Rossi 2009 Table 4 note quotes context) | Conservative default (6,000 pulses/session); no held guideline specifies an absolute daily ceiling. |
| tms.maxPulsesPerWeek |  | pulses | operational default | $>$30,000 pulses/week triggers caution review | None (operational default) | Conservative default (30,000 pulses/week); no held guideline specifies a weekly ceiling. |
| tms.minInterTrainIntervalSec |  | s | literature-derived | $<$5 s triggers caution/unsafe verdict | Rossi 2009, Table 5, p. 2020 (adapted from Chen 1997) | Safety recommendations for inter-train intervals: minimum safe inter-train interval 5 s. |
| tms.conservativeFrequencyHz |  | Hz | literature-anchored | $>$20 Hz triggers caution verdict | Rossi 2009, Table 4, p. 2018 | Rossi 2009 Table 4: safe train duration guidelines verified up to 20 Hz. |
| tms.cautionFrequencyHz |  | Hz | literature-anchored | $>$25 Hz triggers unsafe verdict | Rossi 2009, Table 4, p. 2018 | Rossi 2009 Table 4: 25 Hz caution boundary. |
| tms.maxFrequencyHz |  | Hz | literature-anchored | $>$25 Hz triggers unsafe verdict | Rossi 2009, Table 4, p. 2018 | Rossi 2009 Table 4: 25 Hz maximum tested frequency. |
| tms.conservativeIntensityPctRMT |  | % RMT | literature-anchored | $>$110% RMT triggers caution evaluation | Rossi 2009, Table 4, p. 2018 | Rossi 2009 Table 4: 110% RMT intensity column. |
| tms.cautionIntensityPctRMT |  | % RMT | literature-anchored | $>$120% RMT triggers warning evaluation | Rossi 2009, Table 4, p. 2018 | Rossi 2009 Table 4: 120% RMT intensity column. |
| tms.maxIntensityPctRMT |  | % RMT | literature-anchored | $>$130% RMT triggers unsafe verdict | Rossi 2009, Table 4, p. 2018 | Rossi 2009 Table 4: 130% RMT maximum tested intensity column. |
| tms.conservativePulsesPerTrain |  | pulses | literature-anchored | $>$50 pulses/train triggers caution review | Rossi 2009, § 3.2; Rossi 2021, § 5.1 | Published conventional rTMS protocols typically deliver $\leq$50 pulses per train. Published value, NOT a consensus safety limit. |
| tms.cautionPulsesPerTrain |  | pulses | literature-anchored | $>$80 pulses/train triggers warning review | Rossi 2009, § 3.2; Rossi 2021, § 5.1 | Extended train length (80 pulses) documented in clinical literature. Published value, NOT a consensus safety limit. |
| tms.conservativeTrainDurationSec |  | s | literature-anchored | $>$5 s train duration triggers caution review | Rossi 2009, § 3.2; Rossi 2021, § 5.1 | Conventional train duration boundary (5 s) from standard clinical rTMS protocols. Published value, NOT a consensus safety limit. |
| tms.cautionTrainDurationSec |  | s | literature-anchored | $>$10 s train duration triggers warning review | Rossi 2009, § 3.2; Rossi 2021, § 5.1 | Extended train duration boundary (10 s) from published research trials. Published value, NOT a consensus safety limit. |
| tms.conservativeSessionDurationMin |  | min | operational default | $>$40 min triggers caution review | None (operational default) | Operational session duration guideline (40 min); no guideline states a hard duration cap. |
| tms.cautionSessionDurationMin |  | min | operational default | $>$50 min triggers warning review | None (operational default) | Operational session duration ceiling (50 min); precautionary engineering default. |
| tdcs.commonCurrentRange.min |  | mA | informational | None (non-evaluative advisory context) | Bikson 2016, § 3.1, p. 643 | Lower bound of common clinical current range (1.0 mA); non-evaluative advisory context. |
| tdcs.commonCurrentRange.max |  | mA | literature-anchored | $>$2.0 mA triggers caution verdict | Bikson 2016, § 3.1, p. 643 | Upper bound of common clinical current range (2.0 mA; Bikson 2016). Published value, NOT a consensus safety limit. |
| tdcs.maxCurrentMa |  | mA | literature-derived | $>$4.0 mA triggers unsafe verdict | Chhatbar 2017, Table 1, p. 556; Bikson 2016, p. 643 | Chhatbar 2017: clinical current escalation study up to 4.0 mA without SAEs. |
| tdcs.commonDurationMin.min |  | min | informational | None (non-evaluative advisory context) | Bikson 2016, § 3.1; Antal 2017, § 3.2 | Lower bound of typical session duration (10 min); non-evaluative advisory context. |
| tdcs.commonDurationMin.max |  | min | literature-anchored | $>$30 min triggers caution verdict | Bikson 2016, § 3.1; Antal 2017, § 3.2 | Upper bound of typical clinical session duration (30 min; Bikson 2016). Published value, NOT a consensus safety limit. |
| tdcs.maxSessionDurationMin |  | min | literature-derived | $>$40 min triggers unsafe verdict | Bikson 2016, p. 643; Fregni 2021, § 3 | Bikson 2016: 40 min conventional protocol duration ceiling across 33,000+ sessions without SAE. |
| tdcs.commonCurrentDensityMaPerCm2Max |  | mA/cm^2^ | literature-anchored | $>$0.06 mA/cm^2^ triggers unsafe verdict | Antal 2017, § 3.2, p. 1780 | Published lesion observation point (0.06 mA/cm^2^; Antal 2017) adopted as conservative ceiling. Published value, NOT a consensus safety limit. |
| tdcs.cautionCurrentDensityMaPerCm2Max |  | mA/cm^2^ | literature-anchored | $>$0.06 mA/cm^2^ triggers unsafe verdict (zero-width caution band) | Antal 2017, § 3.2, p. 1780 | Published lesion observation point (0.06 mA/cm^2^; Antal 2017); zero-width caution band triggers unsafe verdict. Published value, NOT a consensus safety limit. |
| tdcs.commonChargeDensityCPerCm2Max |  | C/cm^2^ | operational default | $>$2.4 C/cm^2^ triggers caution verdict | None (operational default) | Conservative charge density precaution (2.4 C/cm^2^); operational default below lesion threshold. |
| tdcs.cautionChargeDensityCPerCm2Max |  | C/cm^2^ | literature-derived | $>$5.24 C/cm^2^ triggers unsafe verdict | Antal 2017, p. 1780 (quoting Liebetanz 2009) | Antal 2017 (quoting Liebetanz 2009): 5.24 C/cm^2^ histologically confirmed rat brain lesion threshold. |
| tdcs.commonWeeklyChargeDensityMax |  | C/cm^2^ | operational default | $>$12.0 C/cm^2^/week triggers caution verdict | None (operational default) | Conservative weekly charge density precaution (12.0 C/cm^2^); operational default. |
| tdcs.cautionWeeklyChargeDensityMax |  | C/cm^2^ | operational default | $>$26.2 C/cm^2^/week triggers unsafe verdict | None (operational default) | Conservative weekly charge density ceiling (26.2 C/cm^2^); operational default. |
| tdcs.conventionalTotalChargeC |  | C | literature-anchored | Exceeding area-normalized equivalent triggers advisory caution | Bikson 2016, p. 643 | Bikson 2016: conventional total charge envelope ceiling (7.2 C). Published value, NOT a consensus safety limit. |
| tdcs.conventionalMaxSessionDurationMin |  | min | informational | None (non-evaluative advisory context) | Bikson 2016, p. 643 | Conventional session duration reference value (40 min; Bikson 2016); non-evaluative advisory context. |
| tdcs.conventionalMaxCurrentMa |  | mA | informational | None (non-evaluative advisory context) | Bikson 2016, p. 643 | Conventional current reference value (4.0 mA; Bikson 2016); non-evaluative advisory context. |
| tdcs.lesionChargeDensityCPerCm2 |  | C/cm^2^ | informational | None (non-evaluative advisory context) | Antal 2017, p. 1780 (Liebetanz 2009) | Reference display threshold for Liebetanz 2009 lesion boundary (5.24 C/cm^2^); non-evaluative advisory context. |
| tdcs.lesionChargeDensityCitation | "Antal 2017, quoting Liebetanz 2009 (rat cortex): "a charge density threshold below 52,400 C/m2" (no histologically detectable brain lesions below it)" | — | informational | None (citation metadata string) | Antal 2017, p. 1780 | Citation metadata string. |
| tdcs.conventionalChargeCitation | "Bikson 2016: "Conventional charge (reflecting duration and intensity) is limited to 7.2 C (e.g. 40 minutes, 3 mA)"" | — | informational | None (citation metadata string) | Bikson 2016, p. 643 | Citation metadata string. |
| pms.conservativeIntensityMax |  | % MSO | operational default | $>$60% MSO triggers caution verdict | None (operational default; Beaulieu 2015 context) | Conservative intensity default ($\leq$60% MSO); operational default. |
| pms.cautionIntensityMax |  | % MSO | operational default | $>$80% MSO triggers warning review | None (operational default) | Caution intensity default ($\leq$80% MSO); operational default. |
| pms.unsafeIntensityAbove |  | % MSO | operational default | $>$90% MSO triggers unsafe verdict | None (operational default) | Unsafe intensity ceiling ($>$90% MSO); operational default. |
| pms.conservativeFrequencyMax |  | Hz | literature-anchored | $>$20 Hz triggers caution verdict | Beaulieu 2015, Table 1, p. 180 | Beaulieu 2015: 20 Hz typical peripheral nerve stimulation frequency. Published value, NOT a consensus safety limit. |
| pms.cautionFrequencyMax |  | Hz | literature-anchored | $>$30 Hz triggers unsafe verdict | Beaulieu 2015, Table 1, p. 180 | Beaulieu 2015: 30 Hz caution frequency for peripheral stimulation. Published value, NOT a consensus safety limit. |
| pms.maxTrainDurationSec |  | s | literature-anchored | $>$10 s triggers unsafe verdict | Beaulieu 2015, Table 1 | Beaulieu 2015: 10 s maximum train duration. Published value, NOT a consensus safety limit. |
| pms.conservativeTrainDurationSec |  | s | literature-anchored | $>$5 s triggers caution verdict | Beaulieu 2015, Table 1 | Beaulieu 2015: 5 s conservative train duration. Published value, NOT a consensus safety limit. |
| pms.maxSessionDurationMin |  | min | operational default | $>$45 min triggers caution verdict | None (operational default) | Maximum session duration (45 min); operational default. |
| tens.minFrequencyHz |  | Hz | literature-anchored | $<$1 Hz triggers caution/unsafe verdict | Vance 2014, § 2, p. 4; Johnson 2021, p. 6 | Vance 2014 & Johnson 2021: minimum effective LF-TENS frequency (1 Hz). |
| tens.maxFrequencyHz |  | Hz | literature-anchored | $>$150 Hz triggers caution/unsafe verdict | Vance 2014, § 2, p. 4; Johnson 2021, p. 6 | Vance 2014 & Johnson 2021: maximum conventional HF-TENS frequency (150 Hz). |
| tens.conservativePulseWidthUs |  | $\mu$s | literature-anchored | $>$250 $\mu$s triggers caution review | Johnson 2021, p. 6 | Johnson 2021: 250 $\mu$s selective A-beta sensory fibre activation threshold. |
| tens.cautionPulseWidthUs |  | $\mu$s | literature-anchored | $>$400 $\mu$s triggers warning/unsafe verdict | Johnson 2021, p. 6 | Johnson 2021: 400 $\mu$s upper range avoiding A-delta/C nociceptive activation. |
| tens.conservativeIntensityMa |  | mA | literature-anchored | $>$30 mA triggers caution verdict | Vance 2014, § 2; Johnson 2022 | Vance 2014: 30 mA typical sensory threshold in clinical TENS. Published value, NOT a consensus safety limit. |
| tens.cautionIntensityMa |  | mA | literature-anchored | $>$50 mA triggers unsafe verdict | Vance 2014, § 2; Johnson 2022 | Vance 2014: 50 mA maximum tolerable sensory intensity. Published value, NOT a consensus safety limit. |
| tens.maxSessionDurationMin |  | min | operational default | $>$120 min triggers caution verdict | None (operational default) | Conservative session duration ceiling (120 min); operational default. |
| tus.conservativeIsppaWcm2 |  | W/cm^2^ | literature-anchored | $>$30 W/cm^2^ triggers caution verdict | Nandi 2024; Pasquinelli 2019, Table 2 | W/cm^2^ conservative baseline from human trial data. Published value, NOT a consensus safety limit. |
| tus.cautionIsppaWcm2 |  | W/cm^2^ | literature-anchored | $>$100 W/cm^2^ triggers warning review | Nandi 2024; Pasquinelli 2019, Table 2 | W/cm^2^ intermediate caution intensity reported across human studies. Published value, NOT a consensus safety limit. |
| tus.unsafeIsppaAbove |  | W/cm^2^ | literature-anchored | $>$200 W/cm^2^ triggers unsafe verdict | Nandi 2024; Pasquinelli 2019, Table 2 | W/cm^2^ upper boundary exceeding common human trial practice. Published value, NOT a consensus safety limit. |
| tus.conservativeDutyCyclePct |  | % | literature-anchored | $>$20% triggers advisory duty-cycle caution | Nandi 2024; Pasquinelli 2019, Table 2 | % duty cycle typical in human neuromodulation trials. Published value, NOT a consensus safety limit. |
| tus.cautionDutyCyclePct |  | % | literature-anchored | $>$50% triggers high duty-cycle warning | Nandi 2024; Pasquinelli 2019, Table 2 | % duty cycle upper empirical boundary (Nandi 2024). Published value, NOT a consensus safety limit. |
| tus.conservativePRFHz |  | Hz | literature-anchored | $>$100 Hz triggers advisory PRF review | Nandi 2024; Pasquinelli 2019, Table 2 | Hz pulse repetition frequency commonly applied in human trials. Published value, NOT a consensus safety limit. |
| tus.cautionPRFHz |  | Hz | literature-anchored | $>$500 Hz triggers high PRF caution review | Nandi 2024; Pasquinelli 2019, Table 2 | Hz high PRF boundary observed in literature. Published value, NOT a consensus safety limit. |
| tus.maxSessionDurationMin |  | min | operational default | $>$20 min triggers caution verdict | None (operational default) | Conservative session duration ceiling (20 min); operational default. |
| tus.fdaDiagnosticIsppaWcm2 |  | W/cm^2^ | informational | None (non-evaluative advisory context) | Pasquinelli 2019, Table 1, p. 3; FDA 2019 | FDA diagnostic ultrasound reference ceiling (190 W/cm^2^); non-evaluative advisory context. |
| tus.isptaInformationalOnly | true | — | informational | None (non-evaluative advisory context) | Aubry 2025, § 2, p. 1898 | ITRUSST consensus excludes ISPTA as a thermal-safety metric; advisory reference flag. |
| tus.miReference |  | — | literature-derived | Diagnostic cavitation baseline (triggers advisory comparison) | Aubry 2025, § 2, p. 1899; FDA 2019 | ITRUSST / FDA: 1.9 diagnostic reference point for cavitation risk. |
| tus.miCautionAbove |  | — | literature-derived | $>$1.9 triggers caution review for cavitation risk | Aubry 2025, § 2, p. 1899; FDA 2019 | ITRUSST / FDA: 1.9 caution review threshold for transcranial mechanical index. |
| tus.tiCautionAbove |  | — | literature-derived | $>$1.5 triggers thermal schedule monitoring | Aubry 2025, § 2, Table 1, p. 1899 | ITRUSST consensus: TI $>$ 1.5 triggers exposure schedule monitoring. |
| tus.tiUnsafeAbove |  | — | literature-derived | $>$6.0 triggers unsafe verdict | Aubry 2025, § 2, Table 1, p. 1899; FDA 2019 | ITRUSST consensus: TI $>$ 6.0 is unacceptable for human transcranial neuromodulation. |
| tus.tiSchedule | Schedule (6 tiers: 80 min–10 s) | s / min | literature-derived | Exceeding TI-indexed time limit triggers unsafe verdict | Aubry 2025, Table 1, p. 1899; Murphy 2025 | ITRUSST consensus thermal index exposure duration schedule. |
| tus.maxTemperatureRiseC |  | ${}^{\circ}$C | literature-derived | $>$2 ${}^{\circ}$C temperature rise triggers unsafe verdict | Aubry 2025, § 2, p. 1899; Murphy 2025 | ITRUSST consensus: $\leq$2 ${}^{\circ}$C cranial temperature rise ceiling. |
| tus.maxAbsoluteTempC |  | ${}^{\circ}$C | literature-derived | $>$39 ${}^{\circ}$C brain temperature triggers unsafe verdict | Aubry 2025, § 2, p. 1899; Murphy 2025 | ITRUSST consensus: 39 ${}^{\circ}$C maximum absolute brain tissue temperature. |
| tus.pulseDurationSafeMaxMs |  | ms | literature-anchored | $>$5 ms pulse duration triggers caution review | Nandi 2024; Legon 2014; Pasquinelli 2019 | ms safe pulse duration ceiling from empirical human trials. Published value, NOT a consensus safety limit. |
| tus.pulseDurationCautionMaxMs |  | ms | literature-anchored | $>$10 ms pulse duration triggers unsafe verdict | Nandi 2024; Legon 2014; Pasquinelli 2019 | ms maximum caution pulse duration boundary. Published value, NOT a consensus safety limit. |
| tus.frequencyMin |  | kHz | literature-anchored | $<$100 kHz triggers caution verdict | Blackmore 2019, § 3, p. 2 | Blackmore 2019: 100 kHz lower bound for transcranial ultrasound transmission. |
| tus.frequencyMax |  | kHz | literature-anchored | $>$1000 kHz triggers caution verdict | Blackmore 2019, § 3, p. 2 | Blackmore 2019: 1000 kHz (1 MHz) upper bound due to skull acoustic attenuation. |
| tus.frequencyOptimalMin |  | kHz | literature-anchored | Below 250 kHz triggers non-optimal focal advisory note | Blackmore 2019; Pasquinelli 2019 | kHz optimal lower frequency for focal transcranial penetration. Published value, NOT a consensus safety limit. |
| tus.frequencyOptimalMax |  | kHz | literature-anchored | Above 700 kHz triggers increased attenuation advisory note | Blackmore 2019; Pasquinelli 2019 | kHz optimal upper frequency balancing attenuation and focal size. Published value, NOT a consensus safety limit. |
| tus.targetDepthWarningMm |  | mm | operational default | $>$80 mm focal depth triggers warning review | None (operational default) | Target depth warning threshold (80 mm); operational default. |
| tus.maxSessionsPerDay |  | sessions/day | operational default | $>$3 sessions/day triggers caution verdict | None (operational default) | Maximum daily sessions cap (3 sessions/day); operational default. |
| vns.transcutaneous.conservativeCurrentMa |  | mA | informational | None (non-evaluative advisory context) | Yap 2020 | Legacy uncalled transcutaneous parameter; non-evaluative. |
| vns.transcutaneous.cautionCurrentMa |  | mA | informational | None (non-evaluative advisory context) | Yap 2020 | Legacy uncalled transcutaneous parameter; non-evaluative. |
| vns.transcutaneous.maxCurrentMa |  | mA | informational | None (non-evaluative advisory context) | Yap 2020 | Legacy uncalled transcutaneous parameter; non-evaluative. |
| vns.transcutaneous.conservativePWUs |  | $\mu$s | informational | None (non-evaluative advisory context) | Yap 2020 | Legacy uncalled transcutaneous parameter; non-evaluative. |
| vns.transcutaneous.cautionPWUs |  | $\mu$s | informational | None (non-evaluative advisory context) | Yap 2020 | Legacy uncalled transcutaneous parameter; non-evaluative. |
| vns.transcutaneous.conservativeFreqHz |  | Hz | informational | None (non-evaluative advisory context) | Yap 2020 | Legacy uncalled transcutaneous parameter; non-evaluative. |
| vns.transcutaneous.cautionFreqHz |  | Hz | informational | None (non-evaluative advisory context) | Yap 2020 | Legacy uncalled transcutaneous parameter; non-evaluative. |
| vns.transcutaneous.maxFreqHz |  | Hz | informational | None (non-evaluative advisory context) | Yap 2020 | Legacy uncalled transcutaneous parameter; non-evaluative. |
| vns.auricular.conservativeCurrentMa |  | mA | literature-anchored | $>$4.0 mA triggers caution verdict | Kim 2022, § 3, p. 4 | Kim 2022 meta-analysis: 4.0 mA standard titration ceiling in taVNS. |
| vns.auricular.cautionCurrentMa |  | mA | literature-anchored | $>$6.0 mA triggers warning review | Kim 2022; Yap 2020 | Kim 2022: 6.0 mA high-intensity titration boundary. Published value, NOT a consensus safety limit. |
| vns.auricular.maxCurrentMa |  | mA | literature-anchored | $>$8.0 mA triggers unsafe verdict | Kim 2022; Yap 2020 | Kim 2022: 8.0 mA upper empirical safety ceiling. Published value, NOT a consensus safety limit. |
| vns.auricular.conservativePWUs |  | $\mu$s | literature-anchored | $>$300 $\mu$s triggers caution verdict | Kim 2022, § 3, p. 4 | Kim 2022 meta-analysis: 300 $\mu$s standard pulse width in 50+ clinical trials. |
| vns.auricular.cautionPWUs |  | $\mu$s | literature-anchored | $>$500 $\mu$s triggers warning review | Kim 2022 | Kim 2022: 500 $\mu$s upper pulse width reported in 25 studies. Published value, NOT a consensus safety limit. |
| vns.auricular.maxPWUs |  | $\mu$s | operational default | $>$1000 $\mu$s triggers unsafe verdict | None (operational default; Kim 2022 context) | Conservative pulse width ceiling (1000 $\mu$s); operational default. |
| vns.auricular.conservativeFreqHz |  | Hz | literature-anchored | $>$25 Hz triggers caution verdict | Kim 2022, § 3, p. 4 | Kim 2022 meta-analysis: 25 Hz standard frequency in 82 clinical trials. |
| vns.auricular.cautionFreqHz |  | Hz | literature-anchored | $>$50 Hz triggers warning review | Kim 2022; Farmer 2021 | Farmer 2021 & Kim 2022: 50 Hz caution frequency boundary. Published value, NOT a consensus safety limit. |
| vns.auricular.maxFreqHz |  | Hz | literature-anchored | $>$100 Hz triggers unsafe verdict | Kim 2022 | Kim 2022: 100 Hz maximum tested frequency in clinical trials. Published value, NOT a consensus safety limit. |
| vns.cervical.conservativeCurrentMa |  | mA | literature-anchored | $>$2.0 mA triggers caution verdict | Redgrave 2018, § 3.2, Table 1, p. 1228 | Redgrave 2018: 2.0 mA conservative cervical current baseline. |
| vns.cervical.cautionCurrentMa |  | mA | literature-anchored | $>$4.0 mA triggers warning review | Redgrave 2018; Yap 2020 | Redgrave 2018: 4.0 mA typical upper titration in cervical nVNS. Published value, NOT a consensus safety limit. |
| vns.cervical.maxCurrentMa |  | mA | literature-anchored | $>$6.0 mA triggers unsafe verdict | Redgrave 2018; Yap 2020 | Redgrave 2018: 6.0 mA maximum cervical current. Published value, NOT a consensus safety limit. |
| vns.cervical.conservativePWUs |  | $\mu$s | literature-anchored | $>$250 $\mu$s triggers caution verdict | Redgrave 2018, § 3.2, Table 1, p. 1228 | Redgrave 2018: 250 $\mu$s standard cervical pulse width. |
| vns.cervical.cautionPWUs |  | $\mu$s | literature-anchored | $>$400 $\mu$s triggers warning review | Redgrave 2018 | Redgrave 2018: 400 $\mu$s caution pulse width. Published value, NOT a consensus safety limit. |
| vns.cervical.maxPWUs |  | $\mu$s | operational default | $>$800 $\mu$s triggers unsafe verdict | None (operational default; Redgrave 2018 context) | Conservative cervical pulse width ceiling (800 $\mu$s); operational default. |
| vns.cervical.conservativeFreqHz |  | Hz | literature-anchored | $>$25 Hz triggers caution verdict | Redgrave 2018, § 3.2, Table 1, p. 1228 | Redgrave 2018: 25 Hz standard cervical stimulation frequency. |
| vns.cervical.cautionFreqHz |  | Hz | literature-anchored | $>$40 Hz triggers warning review | Redgrave 2018; Farmer 2021 | Redgrave 2018 & Farmer 2021: 40 Hz caution frequency. Published value, NOT a consensus safety limit. |
| vns.cervical.maxFreqHz |  | Hz | literature-anchored | $>$80 Hz triggers unsafe verdict | Redgrave 2018 | Redgrave 2018: 80 Hz maximum cervical frequency. Published value, NOT a consensus safety limit. |
