## supplementary-tests-results for "NINM Safety Parameter Explorer: a safety-reference tool for non-invasive neuromodulation"

### **Supplementary Material — S4 - Tests results**

The evaluation of the Explorer comprises two complementary tiers: (1) an external benchmark evaluation examining the engine’s tier-calibrated architecture against five landmark, peer-reviewed, and FDA-cleared neuromodulation protocols; and (2) an internal implementation verification suite measuring self-consistency across 38 curated example protocols, quantified rule coverage, and continuous boundary-value sensitivity. Software quality assurance metrics (Fifteen static guards, headless browser automation, and WCAG 2.1 AA contrast measurements across 401 distinct text styles across both themes, 98 component boundaries, 16 hover states, and 4 focus rings) are detailed in Supplementary Material S3.

To evaluate whether the tier-calibrated architecture prevents spurious safety alarms on established clinical research and regulatory-cleared paradigms, an external benchmark suite was executed across five landmark protocols defined independently in the literature (Table 2): (1) accelerated Stanford Neuromodulation Therapy (SNT/SAINT: 1,800 pulses/session iTBS, 10 sessions/day, 90,000 pulses/5 days [4, 5]); (2) conventional anodal tDCS (2.0 mA on 25 cm² pad, $J=0.080$ mA/cm² [6, 20]); (3) high-definition tDCS (HD-tDCS, 4$\times$1 ring, 2.0 mA on 3.14 cm², $J=0.637$ mA/cm²); (4) gammaCore cervical non-invasive vagus nerve stimulation (FDA 510(k) cleared; 30 mA peak, 1 ms pulse width [28]); and (5) transcranial ultrasound with extended burst duration (30 ms pulse duration, $\text{ISPTA}=1.6$ W/cm^2^ [9, 10]). All five landmark protocols passed evaluation with zero false-unsafe verdicts, correctly triggering calibrated Tier 2 specialist-review caution or Tier 3 operational pacing guidance rather than categorical rejection.

**Table 2:** Benchmark performance across five landmark clinical research and regulatory-cleared neuromodulation protocols ($n=5$). All five protocols evaluated with zero false-unsafe verdicts, correctly triggering calibrated Tier 2 specialist-review caution or Tier 3 operational pacing guidance rather than categorical rejection.

| **Modality** | **Paradigm** | **Independent Source / Clearance** | **Parameter Profile** | **Calibrated Verdict** |
| --- | --- | --- | --- | --- |
| TMS (iTBS) | SNT / SAINT | [4, 5] | 1,800 p/sess, 10 sess/d, 90k p/wk | Caution (Tier 3) |
| tDCS | Conventional Anodal | [6, 20] | 2.0 mA on 25 cm² ($J=0.080$ mA/cm²) | Caution (Tier 2) |
| tDCS | HD-tDCS (4$\times$1 Ring) | Published trial literature | 2.0 mA on 3.14 cm² ($J=0.637$ mA/cm²) | Caution (Tier 2) |
| tVNS | gammaCore nVNS | FDA 510(k) cleared; [28] | 30 mA peak, 1 ms pulse, 2-min cycle | Caution (Tier 2) |
| tUS | Extended Burst tUS | [9, 10] | 30 ms burst, 500 kHz, 1.6 W/cm^2^ | Caution (Tier 2) |

To assert internal implementation consistency, each of the 38 examples in the curated library was loaded into the interface and submitted through the application’s own form-submission path, which is the same path a user takes. For every example, the engine’s computed overall verdict (safe, caution, or unsafe) was compared with the verdict the example library itself assigns to that protocol, fixed at design time when the library was curated. The engine reproduced the assigned verdict in 38 of 38 cases, in every modality and every verdict class (Table 3). Every submission completed without incident: none was rejected by the input checks, none produced an error state, and each returned a definite verdict.

**Table 3:** Engine-vs-declaration agreement on the example-protocol verification suite (simulated evaluation; $n=38$).

| **Modality** | **Agreement** | **Declared bands (safe/caution/unsafe)** |
| --- | --- | --- |
| TMS (incl. TBS) | 12/12 | 6/2/4 |
| tDCS | 5/5 | 2/0/3 |
| PMS | 5/5 | 3/1/1 |
| TENS | 5/5 | 3/1/1 |
| tUS | 5/5 | 1/3/1 |
| tVNS | 6/6 | 4/1/1 |
| **All modalities** | **38/38** | **19/8/11** |

**Table 4:** Provenance-attributed verdict distribution across the 38 verification protocols. Under the tier-calibrated architecture, non-safe protocols are stratified by evidential provenance: Tier 1 consensus safety ceilings enforce hard unsafe stops; Tier 2 empirical trial envelopes trigger specialist review caution; and Tier 3 operational defaults provide pacing guidance.

| **Modality** | **Safe** | **Tier 1 (Unsafe)** | **Tier 2 (Caution)** | **Tier 3 (Pacing Caution)** |
| --- | --- | --- | --- | --- |
| TMS (incl. TBS) | 6 | 4 | 2 | 0 |
| tDCS | 2 | 1 | 2 | 0 |
| PMS | 3 | 1 | 1 | 0 |
| TENS | 3 | 1 | 1 | 0 |
| tUS | 1 | 1 | 3 | 0 |
| tVNS | 4 | 0 | 2 | 0 |
| **All modalities** | **19** | **8** | **11** | **0** |

Under the tier-calibrated verdict architecture (Table 4), non-safe protocol evaluations are stratified according to the evidential tier of the governing rule. High-risk protocols (such as TMS train duration schedule exceedances [16], tUS thermal index exceedances $\text{TI}>6.0$ [10], or tDCS excessive current and duration combinations) are governed by Tier 1 consensus limits and receive an immediate stop (“Exceeds consensus safety limit: not permitted”). Conversely, protocols crossing empirical trial baselines rather than biological hazard limits (such as tDCS current density between 0.057 and 0.200 mA/cm² [20], tUS pulse duration $>5$ ms [9], or cervical tVNS titration bounds [28, 29]) receive a calibrated Tier 2 verdict (“Outside published clinical experience: specialist review”), alerting clinicians to elevated monitoring needs without misbranding research as categorically unsafe. Importantly, for tDCS, the tier-calibrated architecture resolves the previous absence of intermediate caution states, stratifying the five verification protocols into two Safe, two Tier 2 Caution, and one Tier 1 Unsafe.

Rule coverage was quantified across the engine’s 92 evaluative rules (Supplementary Material S3). The 38 verification protocols directly exercise 20 distinct safety rules (21.7% coverage), and 22 rules (23.9%) when combined with the landmark protocol suite. To verify the remaining continuous parameter space, an automated boundary-value testing suite evaluated 10 scalar parameter thresholds across all modalities at $[T-\epsilon,T,T+\epsilon]$ (30 test points; Supplementary Material S3), confirming 100% monotonic safety transitions across critical biological ceilings including tDCS current (4.0 mA), current density (0.057 and 0.200 mA/cm²), charge density (5.24 C/cm²), tUS Thermal Index ($\text{TI}=6.0$), Mechanical Index ($\text{MI}=1.9$), pulse duration (5.0 ms), and TMS Rossi Table 4 train duration limits.

Table 5 reproduces, verbatim from the engine’s output, one verification-suite assessment chosen because its verdict turns on a *derived* quantity rather than on any entered parameter: the entered current (2.00 mA) lies inside the common published range, and only the computed current density crosses a published observation point. This is precisely the arithmetic a laboratory workflow would otherwise perform by hand.

**Table 5:** Example input: output pair (tDCS, DLPFC montage; engine output captured from the pinned build through the real submission path). Citations are the engine’s own attached sources, reproduced as captured [18, 20, 21]; the operational-default label reproduces the engine’s in-app provenance marker attached specifically to unquoted operational defaults (here, the weekly charge cap), while published observation points (the current-density ceiling) carry the literature-anchored non-limit marker.

| **Element** | **Value / finding** |
| --- | --- |
| Inputs | tDCS, anodal, DLPFC; current 2.00 mA; electrode area 25 cm²; duration 30 min; 5 sessions/week |
| Current | mA — safe (below consensus conventional envelope 4 mA [18, 20]; empirical stroke escalation study [21] confirms tolerability at this boundary; common operational range 1–2 mA is an informational reference range, not a consensus safety limit) |
| Current density | $J=I/A=2.00/25.00=0.0800$ mA/cm² — caution (literature-anchored: published value, not a consensus safety limit; ceiling 0.057 mA/cm², adopted from published adverse-event observation [20]; specialist review advised) |
| Charge density | $Q/A=((2.00/1000)\times1800)/25.00=0.144$ C/cm² — safe (evaluated against the rat cortical lesion threshold of 5.24 C/cm² reported by Liebetanz et al. and cited in Antal et al. [20]) |
| Total charge / session | C ($Q=(2.00/1000)\times1800$) — safe (within the $\leq$7.2 C conventional human trial session charge envelope [18]) |
| Session duration | min — safe ($\leq$40 min envelope [18]) |
| Weekly charge density | $Q_{\text{wk}}=5\times0.144=0.72$ C/cm²/wk — safe (conservative weekly limit 12 C/cm²/wk; unsourced operational default) |
| Overall verdict | CAUTION (outside 35 cm² conventional envelope; skin inspection advised; formerly evaluated as UNSAFE PARAMETER PROFILE) |
| Engine warning (verbatim) | “Current density (0.080 mA/cm²) exceeds the published observation point of $\leq$0.06 mA/cm², at which repeated daily tDCS ‘caused persisting skin lesions under the electrodes in some subjects’ (Antal 2017, at 25–35 cm² and 1.5–2.1 mA).” |
| Operational default (verbatim labelling for weekly charge cap) | “UNSOURCED — this is an operational default, not literature-derived: no consensus guideline specifies this value, and it is retained as a precaution rather than presented as cited.” |
