## Supplementary material for "NINM Safety Parameter Explorer: a safety-reference tool for non-invasive neuromodulation": Table 1

| **Modality** | **Literature-derived** | **Literature-anchored** | **Operational default** | **Total** |
| --- | --- | --- | --- | --- |
| TMS | 7 | 4 | 4 | 15 |
| tDCS | 3 | 5 | 3 | 11 |
| PMS | 0 | 4 | 4 | 8 |
| TENS | 4 | 2 | 1 | 7 |
| tUS | 9 | 11 | 3 | 23 |
| tVNS | 6 | 10 | 2 | 18 |
| **All** | **29** | **36** | **17** | **82** |
