## Supplementary material for "NINM Safety Parameter Explorer: a safety-reference tool for non-invasive neuromodulation": Table 2

| **Modality** | **Paradigm** | **Independent Source / Clearance** | **Parameter Profile** | **Calibrated Verdict** |
| --- | --- | --- | --- | --- |
| TMS (iTBS) | SNT / SAINT | [4, 34] | 1,800 p/sess, 10 sess/d, 90k p/wk | Caution (Tier 3) |
| tDCS | Conventional Anodal | [5, 19] | 2.0 mA on 25 cm² ($J=0.080$ mA/cm²) | Caution (Tier 2) |
| tDCS | HD-tDCS (4$\times$1 Ring) | Published trial literature | 2.0 mA on 3.14 cm² ($J=0.637$ mA/cm²) | Caution (Tier 2) |
| tVNS | gammaCore nVNS | FDA 510(k) cleared; [27] | 30 mA peak, 1 ms pulse, 2-min cycle | Caution (Tier 2) |
| tUS | Extended Burst tUS | [8, 9] | 30 ms burst, 500 kHz, 1.6 W/cm^2^ | Caution (Tier 2) |
